# Associations of COVID-19 Symptoms with Omicron Subvariants BA.2 and BA.5, Host Status, and Clinical Outcomes: A Registry-Based Observational Study in Sapporo, Japan

**DOI:** 10.1101/2023.02.02.23285393

**Authors:** Sho Nakakubo, Naoki Kishida, Kenichi Okuda, Keisuke Kamada, Masami Iwama, Masaru Suzuki, Isao Yokota, Yoichi M. Ito, Yasuyuki Nasuhara, Richard C. Boucher, Satoshi Konno

## Abstract

**Background:** Previous SARS-CoV-2 infection and vaccination, coupled to rapid evolution of SARS-CoV-2 variants, have modified COVID-19 clinical manifestations. We characterized clinical symptoms of COVID-19 individuals in omicron BA.2 and BA.5 Japanese pandemic periods to identify omicron and subvariant associations between symptoms, immune status, and clinical outcomes.

**Methods:** Individuals registered in Sapporo’s web-based COVID-19 information system entered 12 pre-selected symptoms, days since symptom onset, vaccination history, SARS-CoV-2 infection history, and background. Symptom frequencies, variables associated with symptoms, and symptoms associated with progression to severe disease were analysed.

**Results:** For all omicron-infected individuals, cough was the most common symptom (62.7%), followed by sore throat (60.7%), nasal discharge (44.3%), and fever (38.8%). Omicron BA.5 infection was associated with a higher symptom burden than BA.2 in vaccinated and unvaccinated individuals. Omicron breakthrough-infected individuals with ≥ 3 vaccinations or previous infection were less likely to exhibit systemic symptoms, but more likely to exhibit upper respiratory symptoms. Infected elderly individuals had lower odds for all symptoms, but, when symptoms were manifest, systemic symptoms were associated with an increased risk, whereas upper respiratory symptoms with a decreased risk, of severe disease.

**Conclusion:** Host immunological status, omicron subvariant, and age were associated with a spectrum of COVID-19 symptoms and outcomes. BA.5 produced a greater symptom burden than BA.2. Vaccination and prior infection mitigated systemic symptoms and improved outcomes, but increased upper respiratory tract symptom burden. Systemic, but not upper respiratory, symptoms in the elderly heralded severe disease.

## Introduction

The coronavirus disease 2019 (COVID-19) pandemic, caused by severe acute respiratory syndrome 2 (SARS-CoV-2), has caused more than 657 million infections worldwide^1^. Since early in the pandemic, multifaceted host factors, e.g., age and underlying disease, have been reported to impact the clinical course of individuals infected with SARS-CoV-2^2-5^. Associated with the widespread implementation of SARS-CoV-2 vaccines and previous infections, the severity and mortality rates of the COVID-19 syndrome have decreased^5-7^. The emergence of mutant strains may have also modified the clinical manifestations of COVID-19^8,9^. A challenge for healthcare providers/public health officials is appropriately advising individuals with breakthrough infections and implementing healthcare strategies in the current omicron pandemic period^10-12^.

Patient clinical symptoms are the most accessible information describing the state of individuals with infections in healthcare settings. However, data linking patient manifestations to viral, host factor, and clinical outcomes must be continuously updated to maximize their utility and guide appropriate healthcare strategies^13,14^. Studies designed to compare the symptoms of alpha vs. delta SARS-CoV-2 variants with those of the original Wuhan strain^15^, and the original omicron variant compared to the delta variant, helped guide healthcare strategies^8^. However, no large-scale epidemiological studies describing COVID-19 symptoms and outcomes as a function of host factors across newer omicron subvariants are available.

Sapporo, a Japanese metropolitan city with 2 million people, launched a registration system to automate the acquisition of self-entered personal information from individuals with COVID-19. This unique system, in which individuals enter their current status via the internet, enables the collection of timely data describing COVID-19 symptoms that can be linked to databases representing individuals’ backgrounds, vaccination and previous SARS-CoV-2 infection status, and severity outcomes. Our objective was to update the characterization of COVID-19 clinical symptoms during the omicron BA.2-BA.5 variant pandemic period and identify associations between clinical symptoms, host factors, immune status, and clinical outcomes relevant to the patient and healthcare communities.

## Methods

### Study population and data sources

This registry-based observational study was based on the Sapporo population. Data from treatment decision sites (TDS) and registration centres for test-positive patients (RCPP) were utilized for analyses (**Fig. S1**). The TDS is a system launched by the City of Sapporo to notify individuals with a COVID-19 diagnosis of the need for hospitalization and clinical advice. Individuals eligible for enrolment in the TDS were: 1) symptomatic individuals who visited a medical institution and tested positive for SARS-CoV-2 (polymerase chain reaction or antigen test), and 2) individuals who were not tested for SARS-CoV-2 but developed new symptoms after a household member tested positive for SARS-CoV-2. Those diagnosed with COVID-19 were requested to immediately register clinical information with the TDS through a web device, including the onset of any newly developed symptoms and the presence/absence of 12 preselected specific symptoms. The RCPP is a registration system operated by the City of Sapporo for symptomatic individuals who cannot visit a healthcare provider. The RCPP sends SARS-CoV-2 antigen test kits to symptomatic individuals at home upon request. A self-test is conducted, and information identical to that for the TDS is entered into the RCPP system by test-positive individuals. Data from both systems are integrated and stored at the Sapporo City Public Health Center. Because both systems share identification information assigned to individuals with COVID-19, case duplication is not possible. Data describing previous SARS-CoV-2 infection and progression to severe disease are collected from the Health Center Real-time Information-sharing System on COVID-19 operated by the Ministry of Health, Labor, and Welfare, as well as data linked to patient information in the TDS and RCPP. Vaccination history, including the timeline of vaccination, was accessed from the Vaccination Record System operated by the Japan Government Digital Agency (**Fig. S1**).

Individuals were excluded from the study if they: (1) entered the date of onset inappropriately, (2) were registered > 5 days after symptom onset, or (3) were asymptomatic. Detailed inclusion and exclusion criteria and the treatment of missing values are described in **Supplementary methods**.

### Data collected

The following data were extracted from the TDS and RCPP: 1) date of first symptom onset; 2) dietary intake; 3) presence of 12 predefined symptoms (fever, cough, sore throat, nasal discharge, sputum, headache, joint or muscle pain, severe fatigue, dyspnoea, diarrhoea, and taste or smell disorder) at registration; and 4) demographic information (age, sex, height, weight, and underlying disease). Outcomes defining severe disease, i.e., requiring oxygenation, mechanical ventilation, or death, were collected as above.

### Characterization of SARS-CoV-2 omicron subvariants

Whole genome analysis was performed at the Sapporo City Institute of Public Health to identify SARS-CoV-2 variants prevalent in Sapporo. Randomly collected specimens with positive SARS-CoV-2 test results were analysed for approximately 50–80 cases per week, and the proportions of omicron subvariants were calculated. In this study, omicron subvariant epidemic periods were defined as when the percentage of the most predominant subvariants exceeded 80%.

### Statistical analysis

Continuous and categorical data were expressed as median (interquartile range [IQR]) and proportions, respectively. A 95% confidence interval (CI) was calculated for symptom frequency, odds ratio (OR), and hazard ratio. Multivariate logistic regression analyses were performed, using days from symptom onset, age, sex, body mass index (BMI), underlying diseases, omicron subvariants, vaccination status, and previous SARS-CoV-2 infection as explanatory variables, with the development of each symptom as the outcome. To estimate symptom time courses, frequencies of the 12 predefined symptoms were calculated from individuals aligned by days from the first symptom onset and plotted on a day 0-5 scale. Estimates of the relative rates of symptom accumulation were analysed using a Cox regression model. Regarding logistic regression analysis for progression to severe disease, all symptoms were incorporated in addition to the factors included in the above-mentioned multivariable symptoms analysis. All statistical analyses were performed using JMP® Pro Version 16.2.0 (SAS Institute Inc., Cary, NC, USA).

## Results

### Study population

Data collected from the TDS and RCPP systems were analysed between April 25, 2022, and September 25, 2022. The epidemic period for the omicron subvariant BA.2 spanned April 25 to June 26, and that for BA.5 spanned July 18 to September 25 (**Fig. 1**). After individuals who met the exclusion criteria were excluded (< 5·6%), a total of 157,861 symptomatic COVID-19 cases (34,336 and 123,525 for the BA.2 and BA.5 groups, respectively) were analysed (**Fig. 1**).

**Figure 1.**
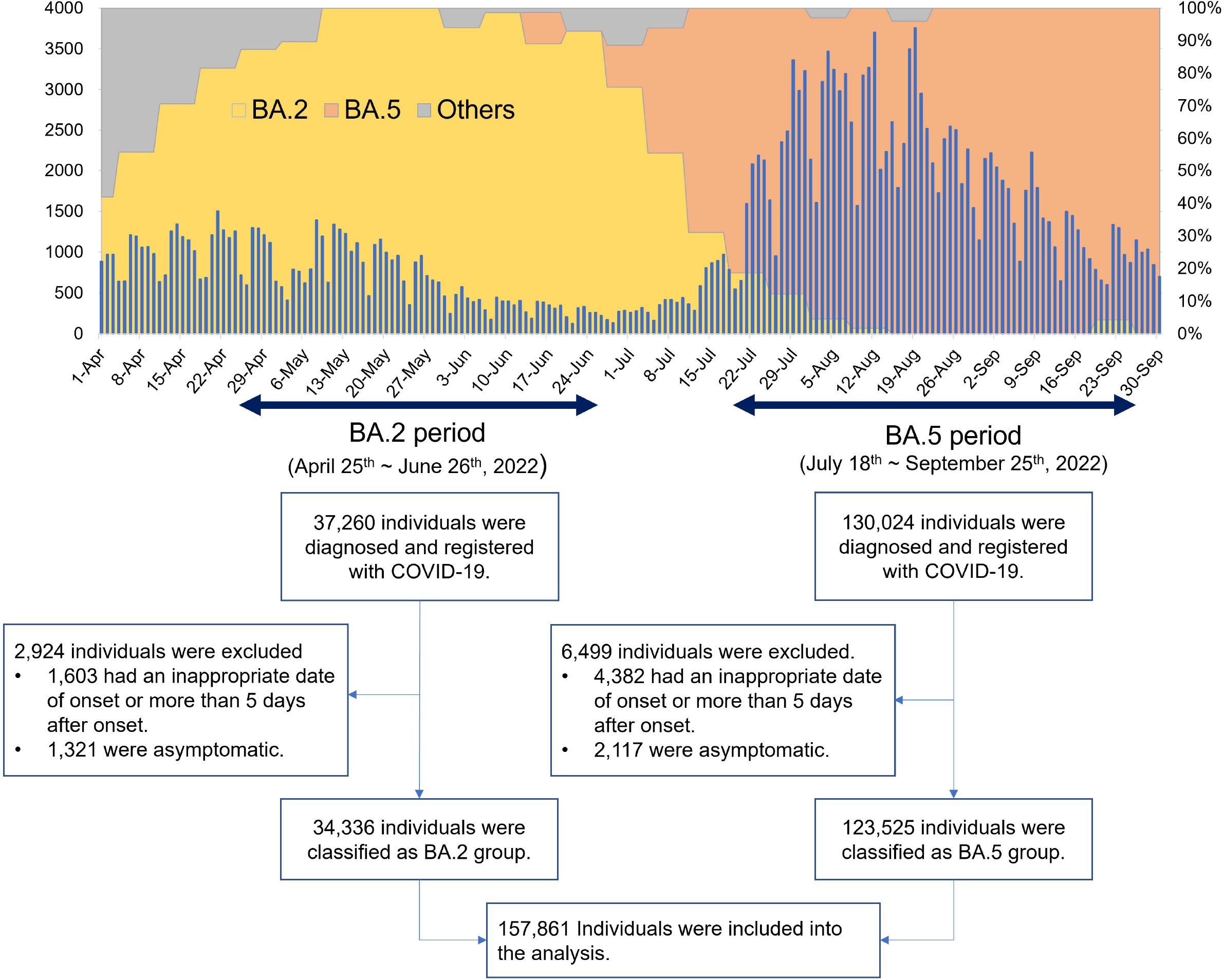
Schematic representations of the study period and eligibility determination. The bars indicate the number of persons with new infections per day in Sapporo. The stacked area graph shows the detection rate of mutant strains in Sapporo. The BA.2 period was defined as when the detection rate of omicron subvariant BA.2 exceeded 80%, and the BA.5 period was defined as when the detection rate of BA.5 exceeded 80% and until the end of notifiable disease surveillance in Sapporo.

The median age of all individuals included in the study was 33 years (range 0–104) (**Table 1**). The most prevalent age group was the 30-year-old group, and those aged ≥ 65 years accounted for 9·3% of the total. Forty-eight percent were men, and the median BMI was 21·1 (IQR 18·6–24·0). The proportion of patients with an underlying disease was highest for hypertension (5·6%), followed by chronic respiratory (3·6%) and cardiovascular diseases (1·0%). SARS-CoV-2 vaccination status was: 38·0% unvaccinated, 0·7% one dose, 24·3% two doses, 33·8% three doses, and 3·2% four doses. Additionally, 3·7% had a history of previous SARS-CoV-2 infection. Coughing was the most prevalent symptom (62·7%), followed by sore throat (60·7%), nasal discharge (44·3%), headache (42·1%), and fever (38·8%) (**Fig. 2A, Table S1**). A total of 142 individuals developed severe disease, and four died within 30 days after onset. The risk of progression to severe disease was 0·02% for individuals younger than 65 years of age and 1·01% for those at this age or over (**Table 1)**.

**Table 1.**
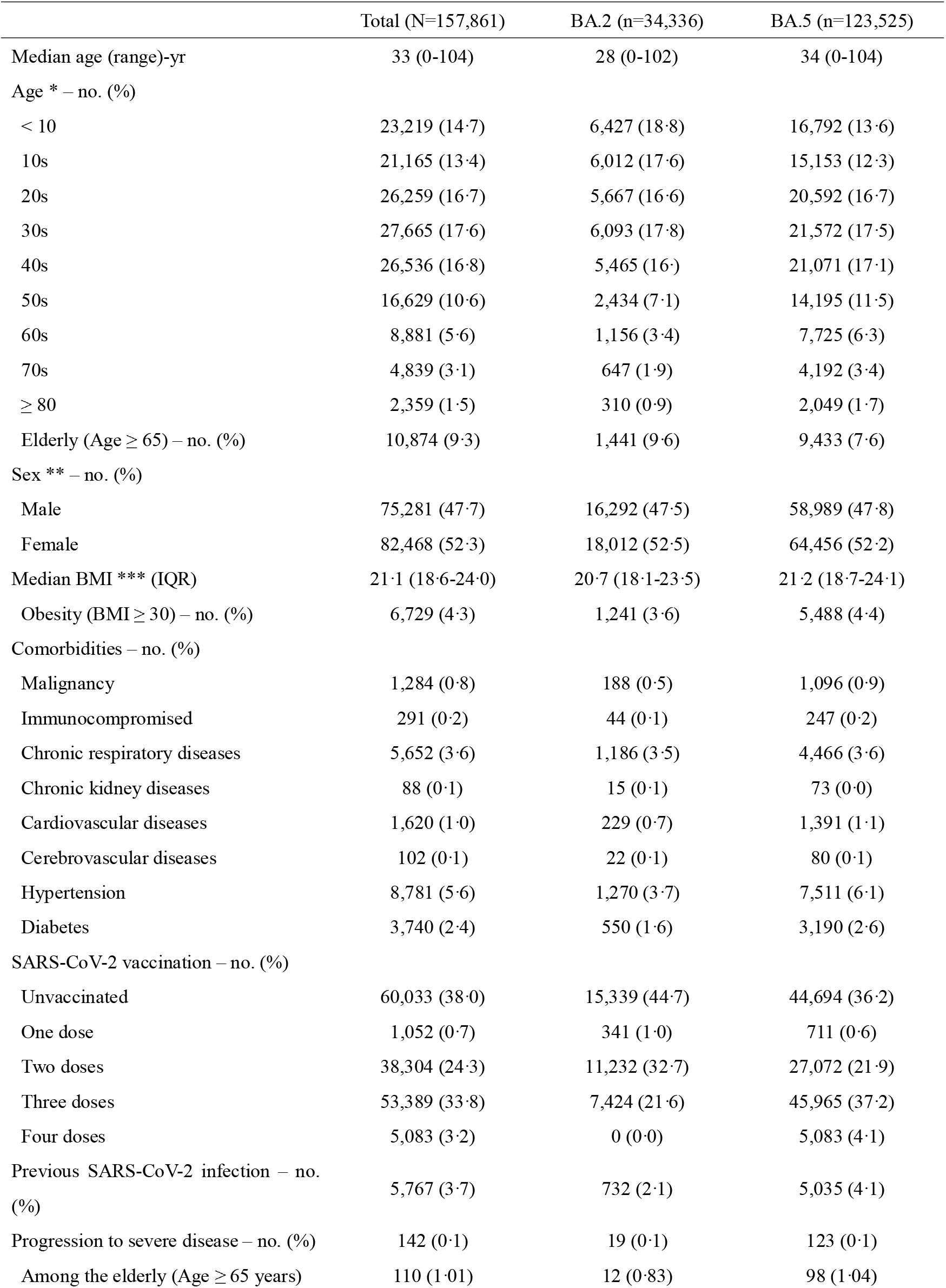
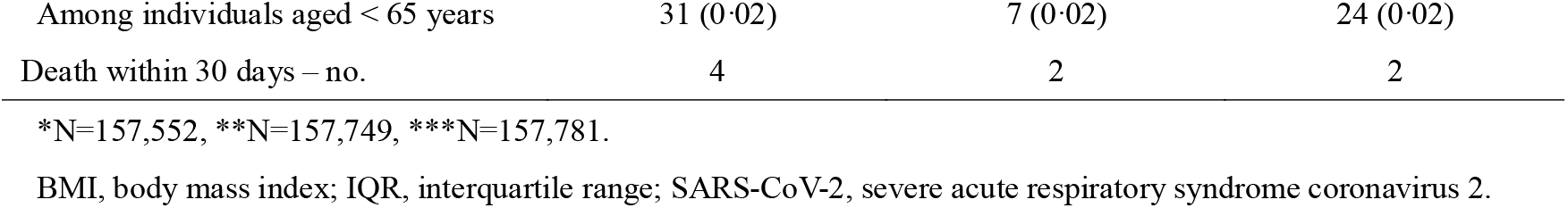
Clinical characteristics of the study population.

**Figure 2.**
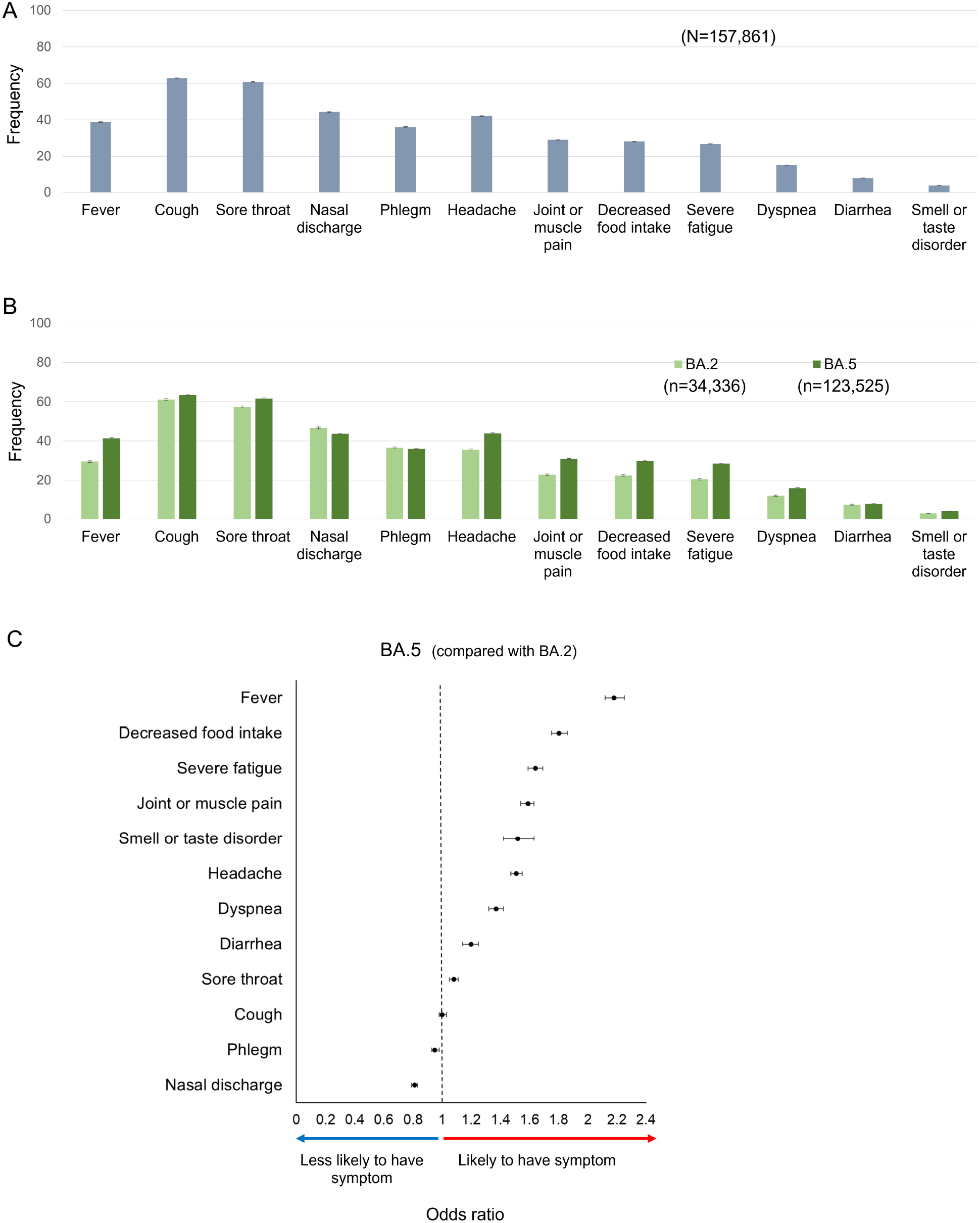
Association of COVID-19 symptoms with the omicron subvariants BA.2 and BA.5. **A**. Frequency of COVID-19 symptoms in the total study population. **B**. Frequency of COVID-19 symptoms in individuals with BA.2 and BA.5 infections. Error bars indicate 95% confidence intervals (CIs). **C**. Associations between omicron subvariants and symptom likelihood. Multivariate analysis used each symptom as an outcome and mutant strain, age, body mass index, underlying disease, vaccination history, and history of spontaneous infection as explanatory variables. The type of omicron subvariant (BA.5 or not) and the adjusted odds ratio (OR) for each symptom are arranged from highest to lowest. Points indicate ORs, and bars indicate 95% CIs. The detailed results of the multivariate analysis are presented in **Table S3**.

### Clinical manifestations of individuals infected with omicron BA.2 or BA.5

The clinical features of symptomatic individuals with BA.2 vs. BA.5 infections were also compared (**Table 1**). The BA.2 group had a lower median age, BMI, history of cerebrovascular disease, and a higher proportion of unvaccinated and ≤ 2 dose-vaccinated individuals. The BA.5 group exhibited higher proportions of other comorbidities, e.g., malignancy, immunodeficiency, cardiovascular disease, hypertension, and diabetes, previous SARS-CoV-2 infection, and contained all individuals with a fourth vaccine dose.

All 12 predefined symptoms, except for nasal discharge and phlegm, were more prevalent in the BA.5 vs. BA.2 populations (**Fig. 2B, Table S1**), findings that were replicated in the unvaccinated BA.5 vs. BA.2 subgroups (**Fig. S2, Table S2**). Multivariate analyses of the BA.5 and BA.2 population data identified associations that predicted an increased risk, for BA.5-infected populations, of fever (adjusted OR [95% CI]: 2·18 [2·12–2·25]), decreased food intake (1·80 [1·75–1·86]), severe fatigue (1·64 [1·59–1·69]), joint or muscle pain, smell or taste disorders, headache, dyspnoea, diarrhoea, and sore throat, juxtaposed to a decreased risk of nasal discharge and phlegm (**Fig. 2C, Table S3)**. Logistic regression analysis demonstrated comparable odds for progression to severe disease between the two subvariants **(Table S4**).

### Associations between vaccine status and COVID-19 symptoms

Individuals were divided into two subgroups to determine the effects of the modification of host immune status by vaccination on the prevalence of COVID-19 symptoms. The boundary between the two subgroups was set at three vaccinations, a number clinically effective for omicron variants^16-18^. The group with ≥ 3 vaccinations exhibited breakthrough infections, and a lower frequency of fever, decreased food intake, severe fatigue, joint or muscle pain, and diarrhoea than the group with ≤ 2 vaccinations (**Fig. 3A, Table S1**). In contrast, cough, sore throat, nasal discharge, and phlegm were more common with breakthrough infections in the ≥ 3 vaccination group.

**Figure 3.**
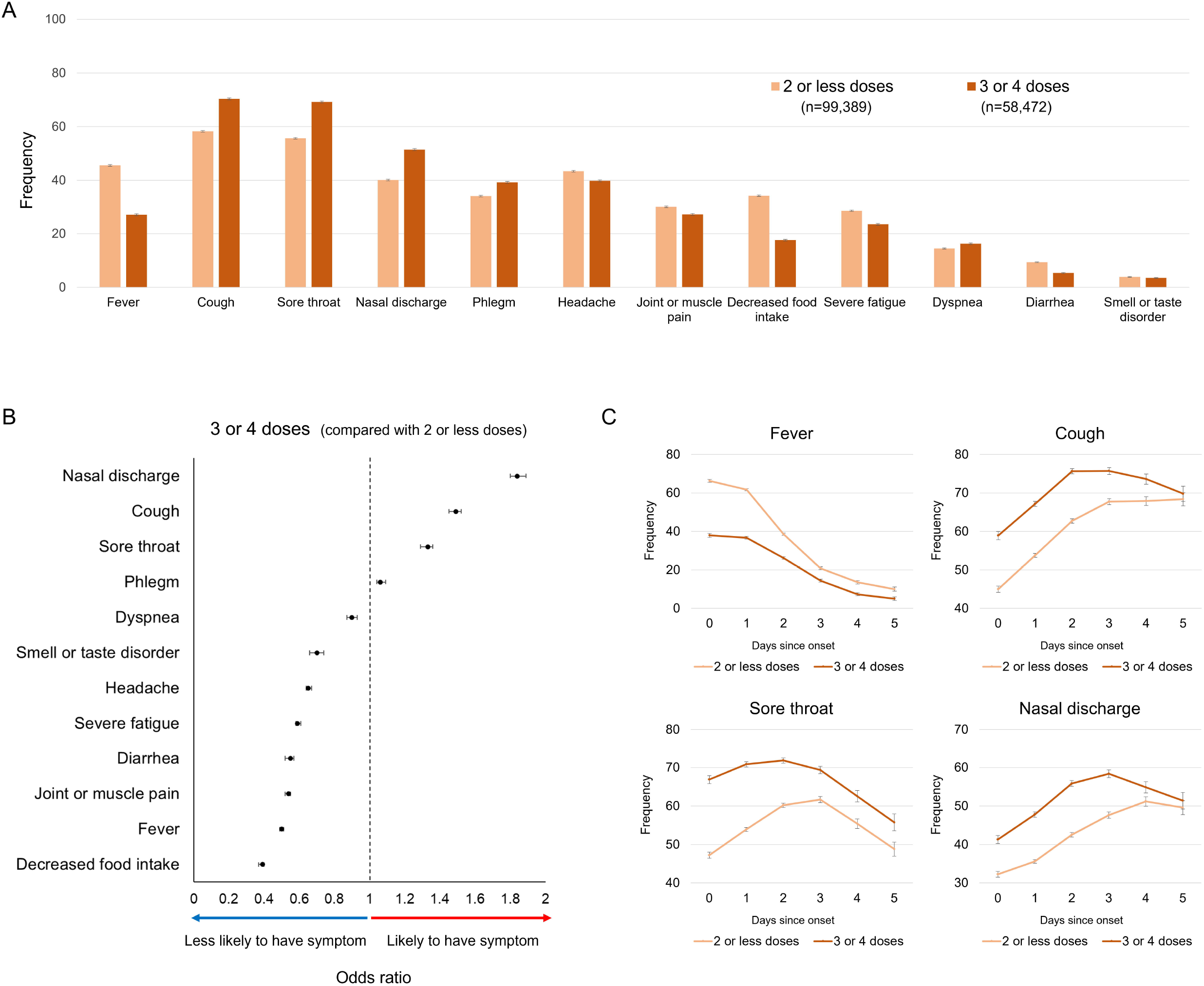
Associations between vaccine status and COVID-19 symptoms. **A**. Symptom frequency is shown in two groups divided by vaccination status. **B**. Associations between vaccination and symptom likelihood. Multivariate analysis used each symptom as an outcome and mutant strain, age, body mass index, underlying disease, vaccination history, and history of spontaneous infection as explanatory variables. The detailed results of the multivariate analysis are presented in **Table S3**. Three or more vaccinations and the adjusted odds ratio (OR) for each symptom are arranged from highest to lowest. Points indicate ORs, and bars indicate 95% confidence intervals (CIs). **C**. Symptom frequency by day since symptom onset in the two groups divided by vaccination status. Error bars indicate 95% CIs.

Logistic regression analysis demonstrated that individuals with breakthrough infections and ≥ 3 vaccinations had a decreased risk of systemic symptoms, including fever (0·50 [0·49–0·51]), decreased food intake (0·39 [0·37–0·40]), severe fatigue (0·59 [0·58–0·61]), joint or muscle pain, headache, diarrhoea, smell or taste disorders, and dyspnoea, as compared to individuals with ≤ 2 vaccinations (**Fig. 3B, Table S3**). In contrast, the likelihood of upper airway symptoms, including nasal discharge (1·84 [1·80–1·89]), cough (1·49 [1·45–1·52]), sore throat (1·33 [1·29–1·36]), and phlegm, were higher in individuals who received ≥ 3 vaccinations than those who received ≤ 2 vaccinations. Similar analyses performed on the ≥ 3 vaccine dose group, incorporating the time elapsed since the last vaccination date as a variable, revealed that longer periods from the last vaccination date were associated with: 1) an increased risk of fever, headache, joint or muscle pain, decreased food intake, and severe fatigue; and 2) a decreased risk of coughing, nasal discharge, sore throat, and dyspnoea (**Table S5**).

The estimated temporal course of breakthrough omicron symptoms differed with vaccination status for upper respiratory but not systemic symptoms (**Fig. 3C, S3, Table S6**). Cox regression analyses, using the presence of symptoms as the occurrence of an event, revealed that the appearance of coughing, sore throat, nasal discharge, and phlegm, but not systemic symptoms, were significantly accelerated in the ≥ 3 vs. ≤ 2 vaccination groups (**Table S7**).

The study population was also divided into subgroups based on a history of previous SARS-CoV-2 infection. The individuals with previous SARS-CoV-2 infection exhibited associations similar to vaccination status, including a higher likelihood of upper respiratory symptoms and a lower likelihood of systemic symptoms (**Fig. S4, Table S3, S6, S7**).

### Associations between COVID-19 symptoms and clinical outcomes in elderly individuals

The frequency of each symptom by age group was calculated (**Fig. 4A, Table S8**). Fever and decreased food intake were most frequent in individuals under 10 years of age and decreased with age. Cough frequency exhibited bimodal peaks in the 20s and 70s age groups. Other symptoms were most frequent in individuals in their 20s and 30s, with frequencies decreasing with increasing age throughout the 70s. Consistent with these data, multivariate analysis identified advanced age (≥ 65 years) as an independent factor associated with a lower likelihood of development of any symptom than younger ages (**Fig. 4B, Table S9**).

**Figure 4.**
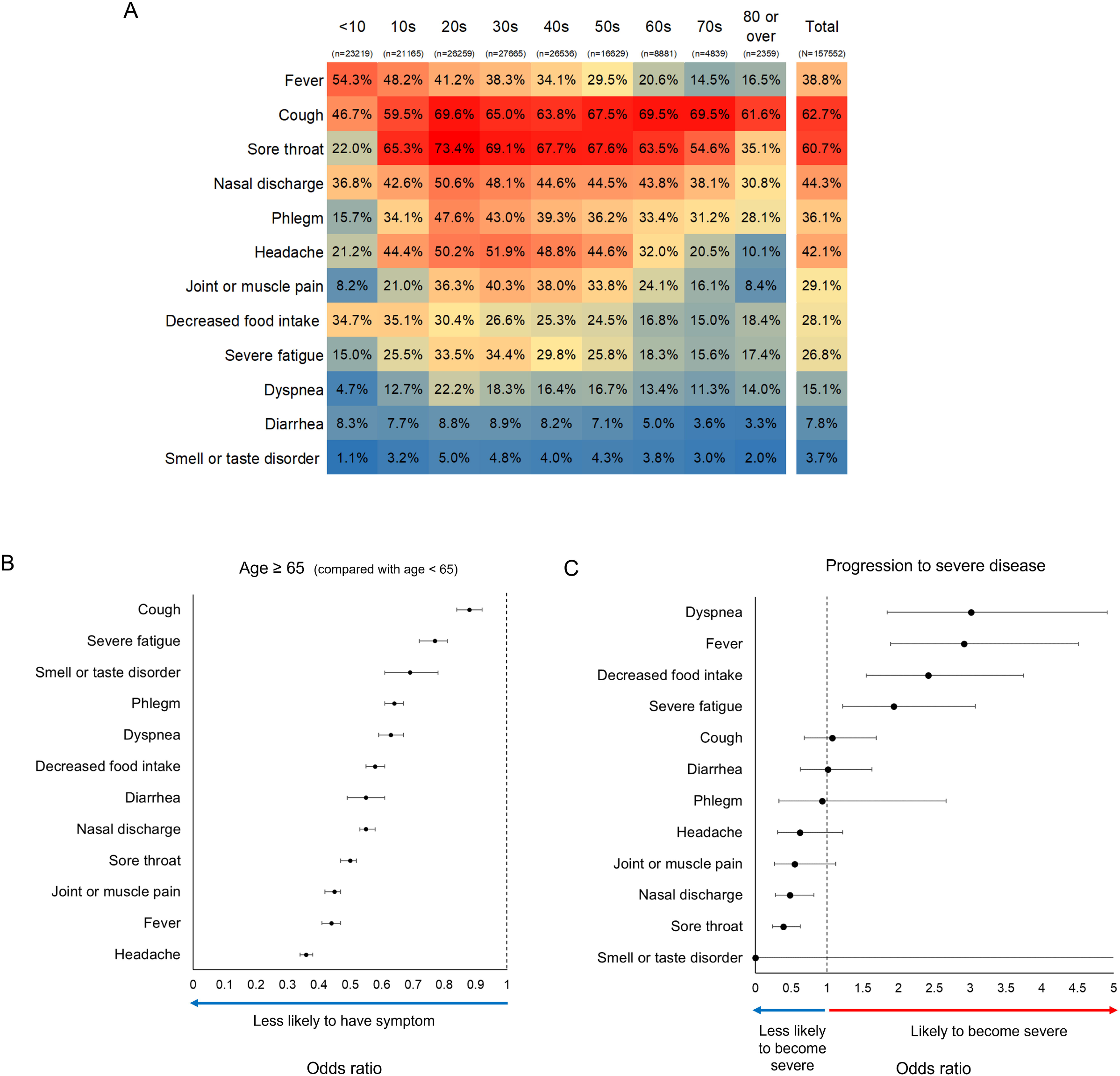
Associations of COVID-19 symptoms with age and progression to severe disease. **A**. Frequency of COVID-19 symptoms according to age. The shade colour of the cells is linked to the high and low percentage values. **B**. Associations between age and symptom likelihood. Multivariate analysis used each symptom as an outcome and mutant strain, age (elderly or non-elderly), body mass index, underlying disease, vaccination history, and history of spontaneous infection as explanatory variables. Elderly individuals (age ≥ 65 years) and the adjusted odds ratio (OR) for each symptom are arranged from highest to lowest. **C**. OR for COVID-19 symptoms and progression to severe disease. Multivariate analysis was performed using progression to severe disease as an outcome and mutant strain, age, body mass index, underlying disease, vaccination history, history of previous infection, and all symptoms as explanatory variables. Symptoms are sorted in descending order of OR for progression to severe disease. Points indicate ORs, and bars indicate 95% confidence intervals. The detailed results of the multivariate analysis are presented in **Table S4**.

Multivariate analyses identified specific COVID-19 symptoms associated with adverse clinical outcomes in elderly individuals. Dyspnoea, fever, decreased food intake, and severe fatigue were associated with an increased risk of severe disease (3·01 [1·84–4·91], 2·91 [1·89–4·51], 2·41 [1·55–3·74], and 1·93 [1·22–3·07], respectively). The combination of these four symptoms was associated with an increased risk of severe disease (adjusted OR [95% CI] for the number of symptoms 1–4 with none as a reference: 2·98 [1·73–5·14], 7·46 [4·15–13·41], 14·38 [7·14–28·98], and 40·72 [14·72–112·68], respectively) (**Fig. 4C, Table S10)**. In contrast, sore throat and nasal discharge were associated with a decreased risk of severe disease (0·39 [0·24–0·63] and 0·48 [0·28–0·82], respectively). The combination of these two upper airway symptoms was associated with a decreased risk of severe disease (adjusted OR [95% CI] with no upper respiratory symptom as a reference: 0·20 [0·09–0·46]) (**Fig. 4C, Table S10**).

## Discussion

The registry-based self-entry COVID-19 symptom study was conducted over an interval when two omicron subvariants were prevalent. Collectively, individuals with omicron infections exhibited more commonly upper respiratory symptoms, e.g., cough, sore throat, and nasal discharge, than systemic symptoms (**Fig. 2A**). The clinical features of individuals with omicron breakthrough infections differed from those in the early Wuhan strain-dominated pandemic period, which was characterized by a higher frequency of fever, cough, dyspnoea, and fatigue and a lower frequency of upper airway symptoms^19-21^. The pattern of a higher incidence of upper respiratory symptoms and a lower incidence of systemic symptoms was replicated in unvaccinated individuals with omicron infections (**Fig. S2**), suggesting that the viral strain is a variable causing difference in clinical manifestations between individuals with the Wuhan strain and omicron breakthrough infection.

The omicron subvariants themselves also differed concerning symptom frequencies. Although emerging later in the pandemic, BA.5 was associated with a higher prevalence of symptoms than BA.2 (**Fig. 2B**). The increased BA.5 symptom prevalence may reflect greater escape from humoral immunity with increased upper airway viral loads as compared to BA.2^22,23^. Notably, the risk of progression to severe disease did not significantly differ between BA.2 vs. BA.5 (**Table S4**).

Significant associations between vaccination status or previous COVID-19 infection and specific symptoms following omicron breakthrough infection were also identified. For example, a reduced risk of systemic symptoms, including fever, fatigue, and headache, was observed in individuals with omicron infections and ≥ 3 vaccinations or a history of previous infection. In contrast, a strong correlation was observed between a history of ≥ 3 vaccinations or previous infection and increased upper airway symptom prevalence (**Fig. 3B, S4**). These contrasting post-vaccination symptom-based observations may reflect: 1) vaccine-mediated reductions in viral load, reduced cytokine release into the systemic circulation, and, hence, reduced systemic symptoms^24-26^, and 2) vaccine-mediated amplification of local host antiviral responses to upper respiratory tract SARS-CoV-2 infection, the consequences being increased symptom prevalence but shorter symptom duration (**Fig. 3**)^27,28^. The increased prevalence of post-breakthrough upper respiratory symptoms after vaccinations likely accounts for the unexplained increase in total symptom burden, but reduced systemic symptoms (fever and chills), recently reported in a cohort of vaccinated United States Essential and Frontline workers after SARS-CoV-2 breakthrough infections^24^.

We observed a decreasing frequency of COVID-19 symptoms with age (**Fig. 4B**). These findings are consistent with an earlier study showing that typical COVID-19 symptoms were less commonly reported in adult groups with advanced age^29,30^. Importantly, if systemic symptoms were present in the elderly (≥ 65-year-old group), strong associations with severe disease were observed (**Fig. 4C**), consistent with reports on pre-vaccinated United States veterans^3^. Unexpectedly, our data suggest that upper respiratory tract symptoms are associated with a reduced risk of severe disease in the elderly.

Our study had several limitations, starting with symptom data being entered directly by individuals without the assistance of healthcare providers. This is a significant limitation as self-reported symptoms are inherently subjective, leading to increased variability. However, these are the data frequently provided in clinical encounters. Second, since symptom data were analysed for 12 predefined questions, COVID-19 symptoms other than the predefined items, including neurological and psychological symptoms, were not evaluated. Third, the reporting of clinical symptom intensity was not collected; thus, quantitative assessments of symptom intensity were not performed. Fourth, asymptomatic individuals were not enrolled in the TDS or RCPP and were excluded from the study. Thus, it is unknown how vaccination and previous infection may have affected the presence of symptoms in the overall population. Fifth, our study was a retrospective observational study and did not provide prospective information describing the duration of COVID-19 symptoms. However, our system, entering symptoms singly at the time of diagnosis with data describing the onset of symptoms, juxtaposed to the large sample size, allowed us to estimate the dynamics of COVID-19 symptoms. Finally, the small number of individuals with severe disease may have resulted in a statistically underpowered detection of factors potentially associated with severe COVID-19 outcomes. Nevertheless, the significance of the specific symptoms identified as independent factors associated with severe outcomes was statistically robust.

In conclusion, our symptom-based description of the clinical manifestations of COVID-19 during the BA.2- and BA.5-dominated COVID-19 pandemic periods provides practical insights into clinical features of the current COVID-19 pandemic. First, BA.5 infections were associated with more prevalent symptoms than BA.2 infections in both vaccinated and unvaccinated individuals. These data suggest that BA.5 emerged as a more troublesome variant than the BA.2 antecedent. Second, an increased prevalence of local upper respiratory tract symptoms, but reduced systemic symptom prevalence, was observed post-vaccination (or previous infection) following omicron breakthrough infections. Thus, it might be appropriate to counsel individuals contemplating vaccination that post-vaccination breakthrough COVID-19 infections may be associated with an increased likelihood of upper airway symptoms, with offsetting benefits being a shorter symptom interval and a reduced risk of severe outcomes. Third, individuals with advanced age experienced, on average, fewer omicron-induced symptoms, but when present, systemic but not upper respiratory symptoms heralded worsened outcomes. These observations may serve as a practical guide to utilize COVID-19 symptoms to predict clinical outcomes for elderly patients with omicron infections.

## Data Availability

The original data is confidential to the City of Sapporo and cannot be made public. All data produced in the present work are contained in the manuscript.

## Acknowledgment

The authors thank the Public Health Office, Health and Welfare Bureau, City of Sapporo, which has long been involved in the identification of all infected populations in Sapporo since the beginning of the COVID-19 epidemic and in the management and integration of the data. Additionally, we thank the Sapporo City Institute of Public Health for providing genomic information on Sapporo’s prevalent SARS-CoV-2 mutant strains.

## Ethical Approval Statement

The research protocol was approved by the Ethics Committee of Hokkaido University Hospital (Research No. 022-0225). Analysis was performed using database in Sapporo city, with no additional risks to the patients. Therefore, the requirement for informed consent from individual participants was waived by the ethics committee. All methods were performed in accordance with the relevant guidelines and regulations of the Ethics Committee of Hokkaido University Hospital. All patient data were anonymized.

## Author contributions

This study was designed by SN, NK, and KO. The underlying data were verified by SN, NK, MI, and YI, and data analyses were performed by SN, YI, and IY. SN wrote the first draft of the manuscript, which was revised by KO, KK, MS, RCB, and KS. All authors interpreted the data, provided a critical review and revision of the text, and approved the final version of the manuscript.

## References

1. WHO COVID-19 Dashboard. Geneva: World Health Organization. Available online: https://covid19whoint/ (last cited: [Jan. 9th, 2023]

2. Terada M, Ohtsu H, Saito S, et al. Risk factors for severity on admission and the disease progression during hospitalisation in a large cohort of patients with COVID-19 in Japan. BMJ Open S 2021; 11(6): e047007.

3. Ioannou GN, Locke E, Green P, et al. Risk Factors for Hospitalization, Mechanical Ventilation, or Death Among 10□131 US Veterans With SARS-CoV-2 Infection. JAMA Netw Open 2020; 3(9): e2022310.

4. O’Driscoll M, Ribeiro Dos Santos G, Wang L, et al. Age-specific mortality and immunity patterns of SARS-CoV-2. Nature 2021; 590(7844): 140–5.

5. Lacy J, Mensah A, Simmons R, et al. Protective effect of a first SARS-CoV-2 infection from reinfection: a matched retrospective cohort study using PCR testing data in England. Epidemiol Infect 2022; 150: e109.

6. Lopez Bernal J, Andrews N, Gower C, et al. Effectiveness of the Pfizer-BioNTech and Oxford-AstraZeneca vaccines on covid-19 related symptoms, hospital admissions, and mortality in older adults in England: test negative case-control study. BMJ 2021; 373: n1088.

7. Shrotri M, Krutikov M, Palmer T, et al. Vaccine effectiveness of the first dose of ChAdOx1 nCoV-19 and BNT162b2 against SARS-CoV-2 infection in residents of long-term care facilities in England (VIVALDI): a prospective cohort study. Lancet Infect Dis 2021; 21(11): 1529–38.

8. Menni C, Valdes AM, Polidori L, et al. Symptom prevalence, duration, and risk of hospital admission in individuals infected with SARS-CoV-2 during periods of omicron and delta variant dominance: a prospective observational study from the ZOE COVID Study. Lancet 2022; 399(10335): 1618–24.

9. Bager P, Wohlfahrt J, Bhatt S, et al. Risk of hospitalisation associated with infection with SARS-CoV-2 omicron variant versus delta variant in Denmark: an observational cohort study. The Lancet Infect Dis 2022; 22(7): 967–76.

10. Dejnirattisai W, Huo J, Zhou D, et al. SARS-CoV-2 Omicron-B.1.1.529 leads to widespread escape from neutralizing antibody responses. Cell 2022; 185(3): 467-84.e15.

11. Martinuzzi E, Boutros J, Glaichenhaus N, Marquette CH, Hofman P, Benzaquen J. Escape of SARS-CoV-2 Variant Omicron to Mucosal Immunity in Vaccinated Subjects. Open Forum Infect Dis 2022; 9(8): ofac362.

12. Kuhlmann C, Mayer CK, Claassen M, et al. Breakthrough infections with SARS-CoV-2 omicron despite mRNA vaccine booster dose. Lancet 2022; 399(10325): 625–6.

13. Alizadehsani R, Alizadeh Sani Z, Behjati M, et al. Risk factors prediction, clinical outcomes, and mortality in COVID-19 patients. J Med Virol 2021; 93(4): 2307–20.

14. Jain V, Yuan JM. Predictive symptoms and comorbidities for severe COVID-19 and intensive care unit admission: a systematic review and meta-analysis. Int J Public Health 2020; 65(5): 533–46.

15. Fernández-de-Las-Peñas C, Cancela-Cilleruelo I, Rodríguez-Jiménez J, et al. Associated-Onset Symptoms and Post-COVID-19 Symptoms in Hospitalized COVID-19 Survivors Infected with Wuhan, Alpha or Delta SARS-CoV-2 Variant. Pathogens 2022; 11(7).

16. Nemet I, Kliker L, Lustig Y, et al. Third BNT162b2 Vaccination Neutralization of SARS-CoV-2 Omicron Infection. N Engl J Med 2022; 386(5): 492–4.

17. Accorsi EK, Britton A, Fleming-Dutra KE, et al. Association Between 3 Doses of mRNA COVID-19 Vaccine and Symptomatic Infection Caused by the SARS-CoV-2 Omicron and Delta Variants. JAMA 2022; 327(7): 639–51.

18. Lauring AS, Tenforde MW, Chappell JD, et al. Clinical severity of, and effectiveness of mRNA vaccines against, covid-19 from omicron, delta, and alpha SARS-CoV-2 variants in the United States: prospective observational study. BMJ 2022; 376: e069761.

19. Komagamine J, Yabuki T. Initial symptoms of patients with coronavirus disease 2019 in Japan: A descriptive study. J Gen Fam Med 2021; 22(1): 61–4.

20. Guan WJ, Ni ZY, Hu Y, et al. Clinical Characteristics of Coronavirus Disease 2019 in China. N Engl J Med 2020; 382(18): 1708–20.

21. Mehta OP, Bhandari P, Raut A, Kacimi SEO, Huy NT. Coronavirus Disease (COVID-19): Comprehensive Review of Clinical Presentation. Front Public Health 2020; 8: 582932.

22. Tuekprakhon A, Nutalai R, Dijokaite-Guraliuc A, et al. Antibody escape of SARS-CoV-2 Omicron BA.4 and BA.5 from vaccine and BA.1 serum. Cell 2022; 185(14): 2422-33.e13.

23. Cao Y, Yisimayi A, Jian F, et al. BA.2.12.1, BA.4 and BA.5 escape antibodies elicited by Omicron infection. Nature 2022; 608(7923): 593–602.

24. Network TH-R. Association of mRNA Vaccination With Clinical and Virologic Features of COVID-19 Among US Essential and Frontline Workers. JAMA 2022; 328(15): 1523–33.

25. Fan Q, Shi J, Yang Y, et al. Clinical characteristics and immune profile alterations in vaccinated individuals with breakthrough Delta SARS-CoV-2 infections. Nat Commun 2022; 13(1): 3979.

26. Mucosal IgA against SARS-CoV-2 Omicron Infection. N Engl J Med 2022; 387(21): e55.

27. Ssemaganda A, Nguyen HM, Nuhu F, et al. Expansion of cytotoxic tissue-resident CD8+ T cells and CCR6+CD161+ CD4+ T cells in the nasal mucosa following mRNA COVID-19 vaccination. Nat Commun 2022; 13(1): 3357.

28. Lim JME, Tan AT, Le Bert N, Hang SK, Low JGH, Bertoletti A. SARS-CoV-2 breakthrough infection in vaccinees induces virus-specific nasal-resident CD8+ and CD4+ T cells of broad specificity. J Exp Med 2022; 219(10).

29. Gómez-Belda AB, Fernández-Garcés M, Mateo-Sanchis E, et al. COVID-19 in older adults: What are the differences with younger patients? Geriatr Gerontol Int 2021; 21(1): 60–5.

30. Trevisan C, Noale M, Prinelli F, et al. Age-Related Changes in Clinical Presentation of Covid-19: the EPICOVID19 Web-Based Survey. Eur J Intern Med 2021; 86: 41–7.

