## Supplementary appendix for "Associations of COVID-19 Symptoms with Omicron Subvariants BA.2 and BA.5, Host Status, and Clinical Outcomes: A Registry-Based Observational Study in Sapporo, Japan"

**The COVID-19 syndrome in the omicron era: symptoms, vaccination status, and clinical outcomes**

Supplementary methods	P2

Table S1. Frequency of COVID-19 symptoms in total and subgroups	P3

Table S2. COVID-19 symptoms frequency in unvaccinated individuals and subgroups	P4

Table S3. Logistic regression analysis of factors for COVID-19 symptoms	P5

Table S4. Logistic regression analysis of factors for severe disease progression	P9

Table S5. Logistic regression analysis of factors for symptoms in the group with three or more vaccination	P10

Table S6. COVID-19 symptoms frequency by days since symptom onset in total and subgroups	P14

Table S7. Cox regression analysis for presence of symptoms	P19

Table S8. Frequency of COVID-19 symptoms by age	P22

Table S9. Logistic regression analysis of factors for symptoms (set elderly as explanatory variable)	P23

Table S10. Logistic regression analysis of factors (including symptom scores) for progression to severe disease	P25

Figure S1. Schema of information registration system for symptomatic COVID-19 patients in Sapporo	P26

Figure S2. COVID-19 symptom frequency in unvaccinated individuals	P27

Figure S3. Frequency of COVID-19 symptoms by days since symptom onset	P28

Figure S4. Association between history of previous infection and COVID-19 symptoms	P29

**Supplementary Methods**

**Inclusion criteria**

Individuals diagnosed with COVID-19 and whose information was registered on the Treatment Decision Site or the Registration Center for Test Positive Patients website during the study periods.

**Exclusion criteria**

1) Incorrect input value for the date of onset (later than the registration date).

2) Registration date is more than five days after the onset date.

3) Under the term "Symptoms to date", "Nothing" was selected. Applicable cases were treated as asymptomatic individuals and excluded.

**Definition of symptoms**

Fever was defined as body temperature ≥ 38 °C.

“Decreased food intake” was defined as less than 30% of baseline food intake.

**Definition of missing values and treatment**

The missing values were applicable for age, sex, and body mass index (BMI). For age, values above 120 were determined to be erroneously entered and were considered missing values. Missing values were assigned to cases in which sex was unknown. Several researchers determined BMI to be an inappropriate value by comparing height and weight with age. Inappropriate values were considered as missing values. Finally, a BMI of < 8 and > 70 were determined to be inappropriate values. The number and frequency of cases in each category were calculated, excluding cases with missing values. Multivariate analysis was performed for cases without missing values.

**Figure drawing**

Microsoft Excel^®^ (Microsoft Corporation, Redmond, WA, USA) was used to draw bar graphs, tables with color scales, and forest plots.

**Table S1. Frequency of COVID-19 symptoms in total and subgroups**

|  | Total | Subvariants | | Vaccination doses | | Previous SARS-CoV-2 infection | |
| --- | --- | --- | --- | --- | --- | --- | --- |
|  | (N=157,861) | BA.2  (n=34,336) | BA.5  (n=123,525) | 2 or less  (n=99,389) | 3 or more  (n=58,472) | No previous infection  (n=152,094) | Previous infection  (n=5,797) |
|  | Frequency - % (95% CI) | | | | | | |
| Fever | 38.8 (38.5 - 39.0) | 29.5 (29.0 - 30.0) | 41.4 (41.1 - 41.6) | 45.6 (45.3 - 45.9) | 27.2 (26.9 - 27.6) | 39.1 (38.9 - 39.4) | 29.3 (28.2 - 30.5) |
| Cough | 62.7 (62.5 - 63.0) | 61.1 (60.5 - 61.6) | 63.2 (62.9 - 63.5) | 58.3 (58.0 - 58.6) | 70.3 (70.0 - 70.7) | 62.7 (62.5 - 63.0) | 62.4 (61.1 - 63.6) |
| Sore throat | 60.7 (60.5 - 61.0) | 57.3 (56.8 - 57.8) | 61.7 (61.4 - 61.9) | 55.7 (55.4 - 56.0) | 69.2 (68.9 - 69.6) | 60.8 (60.5 - 61.0) | 58.7 (57.4 - 59.9) |
| Nasal discharge | 44.3 (44.1 - 44.6) | 46.7 (46.2 - 47.3) | 43.7 (43.4 - 43.9) | 40.1 (39.8 - 40.4) | 51.5 (51.0 - 51.9) | 44.1 (43.8 - 44.3) | 51.4 (50.1 - 52.7) |
| Phlegm | 36.1 (35.8 - 36.3) | 36.4 (35.9 - 37.0) | 35.9 (35.7 - 36.2) | 34.2 (33.9 - 34.5) | 39.3 (38.9 - 39.7) | 36.2 (36.0 - 36.4) | 32.1 (30.9 - 33.3) |
| Headache | 42.1 (41.8 - 42.3) | 35.6 (35.1 - 36.1) | 43.9 (43.6 - 44.2) | 43.4 (43.1 - 43.7) | 39.8 (39.4 - 40.2) | 42.2 (42.0 - 42.5) | 37.7 (36.5 - 39.0) |
| Joint or muscle pain | 29.1 (28.9 - 29.3) | 22.8 (22.4 - 23.2) | 30.9 (30.6 - 31.1) | 30.2 (29.9 - 30.5) | 27.3 (27.0 - 27.7) | 29.4 (29.2 - 29.6) | 21.2 (20.2 - 22.3) |
| Decreased food intake | 28.1 (27.9 - 28.3) | 22.4 (21.9 - 22.8) | 29.7 (29.4 - 29.9) | 34.3 (34.0 - 34.6) | 17.6 (17.3 - 17.9) | 28.4 (28.2 - 28.7) | 19.6 (18.6 - 20.6) |
| Severe fatigue | 26.8 (26.5 - 27.0) | 20.6 (20.1 - 21.0) | 28.5 (28.2 - 28.7) | 28.7 (28.4 - 29.0) | 23.5 (23.1 - 23.8) | 27.0 (26.7 - 27.2) | 21.2 (20.2 - 22.3) |
| Dyspnea | 15.1 (15.0 - 15.3) | 12.1 (11.8 - 12.4) | 16.0 (15.8 - 16.2) | 14.5 (14.3 - 14.7) | 16.3 (16.0 - 16.6) | 15.1 (14.9 - 15.3) | 15.7 (14.8 - 16.7) |
| Diarrhea | 7.9 (7.7 - 8.0) | 7.5 (7.3 - 7.8) | 7.9 (7.8 - 8.1) | 9.3 (9.1 - 9.5) | 5.3 (5.2 - 5.5) | 7.8 (7.7 - 8.0) | 8.2 (7.5 - 8.9) |
| Smell or taste disorder | 3.7 (3.7 - 3.8) | 2.8 (2.6 - 3.0) | 4.0 (3.9 - 4.1) | 3.9 (3.8 - 4.0) | 3.5 (3.4 - 3.7) | 3.8 (3.7 - 3.9) | 3.3 (2.9 - 3.8) |

CI, confidence intervals; SARS-CoV-2, severe acute respiratory syndrome coronavirus 2.

**Table S2. COVID-19 symptoms frequency in unvaccinated individuals and subgroups**

|  | Total | Subvariants | |
| --- | --- | --- | --- |
|  | (N=60,033) | BA.2  (n=15,339) | BA.5  (n=44,694) |
|  | Frequency - % (95% CI) | | |
| Fever | 48.9 (48.5 - 49.2) | 38.2 (37.4 - 38.9) | 52.5 (52.1 - 53.0) |
| Cough | 52.9 (52.5 - 53.3) | 52.0 (51.2 - 52.8) | 53.3 (52.8 - 53.7) |
| Sore throat | 47.3 (46.9 - 47.7) | 43.2 (42.4 - 44.0) | 48.7 (48.2 - 49.1) |
| Nasal discharge | 35.6 (35.3 - 36.0) | 38.3 (37.6 - 39.1) | 34.7 (34.3 - 35.2) |
| Phlegm | 29.3 (29.0 - 29.7) | 29.2 (28.5 - 30.0) | 29.3 (28.9 - 29.8) |
| Headache | 39.6 (39.2 - 40.0) | 32.8 (32.1 - 33.6) | 41.9 (41.4 - 42.3) |
| Joint or muscle pain | 26.4 (26.0 - 26.7) | 20.4 (19.7 - 21.0) | 28.4 (28.0 - 28.9) |
| Decreased food intake | 37.4 (37.0 - 37.8) | 29.3 (28.6 - 30.0) | 40.2 (39.8 - 40.7) |
| Severe fatigue | 27.0 (26.6 - 27.3) | 20.2 (19.6 - 20.9) | 29.3 (28.9 - 29.7) |
| Dyspnea | 12.0 (11.7 - 12.2) | 9.1 (8.7 - 9.6) | 13.0 (12.7 - 13.3) |
| Diarrhea | 9.9 (9.7 - 10.1) | 9.2 (8.8 - 9.7) | 10.1 (9.9 - 10.4) |
| Smell or taste disorder | 3.7 (3.5 - 3.8) | 2.5 (2.3 - 2.8) | 4.1 (3.9 - 4.3) |

CI, confidence intervals; SARS-CoV-2, severe acute respiratory syndrome coronavirus 2.

**Table S3. Logistic regression analysis of factors for COVID-19 symptoms**

A. Fever, cough, sore throat, nasal discharge, phlegm, headache

|  | Fever | Cough | Sore throat | Nasal discharge | Phlegm | Headache |
| --- | --- | --- | --- | --- | --- | --- |
|  | Adjusted OR (95% CI) | | | | | |
| Days from onset |  | | | | | |
| 0 | (reference) | (reference) | (reference) | (reference) | (reference) | (reference) |
| 1 | 0.85 (0.82 - 0.88) | 1.40 (1.36 - 1.45) | 1.19 (1.15 - 1.23) | 1.21 (1.17 - 1.25) | 1.34 (1.29 - 1.39) | 1.00 (0.96 - 1.03) |
| 2 | 0.39 (0.38 - 0.41) | 2.00 (1.93 - 2.06) | 1.33 (1.29 - 1.38) | 1.63 (1.58 - 1.69) | 1.92 (1.85 - 1.99) | 0.79 (0.77 - 0.82) |
| 3 | 0.17 (0.16 - 0.18) | 2.29 (2.20 - 2.38) | 1.28 (1.23 - 1.33) | 1.95 (1.88 - 2.03) | 2.16 (2.07 - 2.25) | 0.57 (0.55 - 0.59) |
| 4 | 0.10 (0.09 - 0.10) | 2.20 (2.10 - 2.32) | 0.96 (0.91 - 1.01) | 2.04 (1.94 - 2.13) | 2.13 (2.02 - 2.23) | 0.43 (0.41 - 0.45) |
| 5 | 0.07 (0.06 - 0.08) | 2.07 (1.94 - 2.22) | 0.73 (0.69 - 0.78) | 1.88 (1.76 - 2.00) | 2.11 (1.98 - 2.26) | 0.35 (0.33 - 0.38) |
| BA.5 group  (compared with BA.2) | 2.18 (2.12 - 2.25) | 1.00 (0.98 - 1.03) | 1.08 (1.05 - 1.11) | 0.81 (0.79 - 0.83) | 0.95 (0.93 - 0.98) | 1.51 (1.47 - 1.55) |
| Age |  |  |  |  |  |  |
| <10 | (reference) | (reference) | (reference) | (reference) | (reference) | (reference) |
| 10s | 0.94 (0.90 - 0.98) | 1.52 (1.46 - 1.58) | 6.49 (6.22 - 6.77) | 1.13 (1.08 - 1.17) | 2.63 (2.51 - 2.75) | 3.35 (3.21 - 3.50) |
| 20s | 0.82 (0.78 - 0.85) | 2.12 (2.04 - 2.20) | 8.91 (8.54 - 9.30) | 1.35 (1.30 - 1.40) | 4.43 (4.24 - 4.63) | 4.61 (4.43 - 4.81) |
| 30s | 0.74 (0.71 - 0.77) | 1.64 (1.58 - 1.70) | 7.04 (6.76 - 7.35) | 1.14 (1.10 - 1.19) | 3.61 (3.46 - 3.78) | 5.06 (4.85 - 5.27) |
| 40s | 0.64 (0.61 - 0.66) | 1.47 (1.42 - 1.53) | 6.46 (6.19 - 6.74) | 0.93 (0.89 - 0.96) | 3.02 (2.88 - 3.16) | 4.61 (4.42 - 4.81) |
| 50s | 0.53 (0.50 - 0.55) | 1.62 (1.55 - 1.69) | 6.15 (5.85 - 6.45) | 0.85 (0.81 - 0.89) | 2.59 (2.46 - 2.72) | 4.09 (3.89 - 4.29) |
| 60s | 0.37 (0.35 - 0.39) | 1.61 (1.52 - 1.70) | 4.95 (4.67 - 5.25) | 0.73 (0.69 - 0.77) | 2.18 (2.05 - 2.32) | 2.65 (2.50 - 2.82) |
| 70s | 0.26 (0.24 - 0.29) | 1.49 (1.39 - 1.61) | 3.38 (3.15 - 3.63) | 0.53 (0.49 - 0.57) | 1.88 (1.74 - 2.03) | 1.55 (1.43 - 1.69) |
| ≥80 | 0.24 (0.22 - 0.28) | 1.11 (1.01 - 1.22) | 1.49 (1.36 - 1.65) | 0.40 (0.36 - 0.44) | 1.72 (1.55 - 1.91) | 0.60 (0.52 - 0.70) |
| Male | 1.14 (1.11 - 1.16) | 0.85 (0.83 - 0.87) | 0.84 (0.82 - 0.86) | 0.90 (0.88 - 0.92) | 0.96 (0.94 - 0.98) | 0.72 (0.70 - 0.73) |
| Obesity (BMI≥30) | 1.00 (0.95 - 1.06) | 1.38 (1.31 - 1.46) | 0.90 (0.85 - 0.95) | 1.50 (1.43 - 1.58) | 1.37 (1.30 - 1.44) | 1.02 (0.97 - 1.07) |
| Comorbidities |  |  |  |  |  |  |
| Malignancy | 1.47 (1.27 - 1.70) | 1.11 (0.97 - 1.27) | 0.99 (0.87 - 1.13) | 0.99 (0.87 - 1.13) | 1.10 (0.97 - 1.25) | 0.99 (0.86 - 1.13) |
| Immuno-compromised | 1.00 (0.74 - 1.34) | 0.87 (0.66 - 1.14) | 0.95 (0.73 - 1.24) | 1.05 (0.81 - 1.36) | 1.21 (0.93 - 1.57) | 1.18 (0.89 - 1.55) |
| Chronic respiratory diseases | 1.04 (0.98 - 1.10) | 1.47 (1.39 - 1.56) | 0.95 (0.90 - 1.01) | 1.41 (1.33 - 1.48) | 1.60 (1.52 - 1.69) | 1.36 (1.28 - 1.43) |
| Chronic kidney diseases | 0.82 (0.48 - 1.39) | 0.81 (0.52 - 1.24) | 0.80 (0.52 - 1.24) | 1.22 (0.79 - 1.88) | 0.79 (0.48 - 1.29) | 0.56 (0.32 - 1.00) |
| Cardiovascular diseases | 1.19 (1.05 - 1.35) | 1.22 (1.09 - 1.36) | 1.14 (1.03 - 1.27) | 1.17 (1.05 - 1.30) | 1.16 (1.04 - 1.29) | 0.94 (0.83 - 1.06) |
| Cerebrovascular diseases | 1.18 (0.71 - 1.96) | 0.92 (0.61 - 1.40) | 0.92 (0.62 - 1.39) | 1.03 (0.69 - 1.55) | 0.87 (0.57 - 1.34) | 0.63 (0.37 - 1.08) |
| Hypertension | 1.04 (0.98 - 1.10) | 1.12 (1.07 - 1.19) | 1.00 (0.95 - 1.06) | 1.09 (1.03 - 1.14) | 1.08 (1.03 - 1.14) | 0.88 (0.83 - 0.93) |
| Diabetes | 0.88 (0.81 - 0.96) | 1.12 (1.04 - 1.21) | 1.00 (0.93 - 1.07) | 1.14 (1.06 - 1.22) | 1.04 (0.96 - 1.11) | 0.91 (0.84 - 0.99) |
| Vaccination ≥3 doses | 0.50 (0.49 - 0.51) | 1.49 (1.45 - 1.52) | 1.33 (1.29 - 1.36) | 1.84 (1.80 - 1.89) | 1.06 (1.04 - 1.09) | 0.65 (0.64 - 0.67) |
| Previous SARS-CoV-2 infection | 0.41 (0.39 - 0.44) | 1.13 (1.07 - 1.20) | 1.10 (1.03 - 1.16) | 1.50 (1.42 - 1.59) | 0.87 (0.82 - 0.92) | 0.75 (0.71 - 0.80) |

BMI, body mass index; CI, confidence interval; OR, odds ratio; SARS-CoV-2, severe acute respiratory syndrome coronavirus 2.

B. Joint or muscle pain, decreased food intake, severe fatigue, dyspnea, diarrhea, smell or taste disorder

|  | Joint or muscle pain | Decreased food intake | Severe fatigue | Dyspnea | Diarrhea | Smell or taste disorder |
| --- | --- | --- | --- | --- | --- | --- |
|  | Adjusted OR (95% CI) | | | | | |
| Days from onset |  | | | | | |
| 0 | (reference) | (reference) | (reference) | (reference) | (reference) | (reference) |
| 1 | 1.08 (1.05 - 1.12) | 1.03 (0.99 - 1.06) | 0.94 (0.90 - 0.97) | 1.09 (1.04 - 1.14) | 1.28 (1.20 - 1.37) | 1.03 (0.93 - 1.14) |
| 2 | 0.83 (0.80 - 0.86) | 0.82 (0.79 - 0.85) | 0.69 (0.66 - 0.71) | 1.28 (1.22 - 1.34) | 1.60 (1.50 - 1.71) | 1.48 (1.34 - 1.63) |
| 3 | 0.57 (0.55 - 0.60) | 0.59 (0.57 - 0.62) | 0.50 (0.48 - 0.52) | 1.31 (1.24 - 1.38) | 1.64 (1.52 - 1.76) | 2.04 (1.84 - 2.27) |
| 4 | 0.39 (0.37 - 0.42) | 0.45 (0.42 - 0.48) | 0.40 (0.37 - 0.42) | 1.27 (1.19 - 1.36) | 1.59 (1.46 - 1.74) | 2.49 (2.21 - 2.80) |
| 5 | 0.30 (0.28 - 0.33) | 0.37 (0.34 - 0.40) | 0.35 (0.32 - 0.38) | 1.19 (1.09 - 1.30) | 1.58 (1.41 - 1.78) | 4.02 (3.53 - 4.57) |
| BA.5 group  (compared with BA.2) | 1.59 (1.54 - 1.63) | 1.80 (1.75 - 1.86) | 1.64 (1.59 - 1.69) | 1.37 (1.32 - 1.42) | 1.20 (1.14 - 1.25) | 1.52 (1.42 - 1.63) |
| Age |  |  |  |  |  |  |
| <10 | (reference) | (reference) | (reference) | (reference) | (reference) | (reference) |
| 10s | 3.35 (3.17 - 3.55) | 1.18 (1.13 - 1.22) | 2.19 (2.09 - 2.30) | 2.94 (2.73 - 3.16) | 0.94 (0.87 - 1.00) | 2.96 (2.56 - 3.43) |
| 20s | 8.34 (7.89 - 8.80) | 1.10 (1.06 - 1.15) | 3.65 (3.49 - 3.82) | 5.66 (5.28 - 6.06) | 1.17 (1.10 - 1.25) | 4.66 (4.06 - 5.35) |
| 30s | 10.40 (9.85 - 10.98) | 0.96 (0.92 - 1.00) | 3.93 (3.76 - 4.12) | 4.38 (4.09 - 4.70) | 1.23 (1.15 - 1.31) | 4.52 (3.94 - 5.19) |
| 40s | 9.81 (9.28 - 10.37) | 0.96 (0.92 - 1.00) | 3.27 (3.12 - 3.42) | 3.74 (3.48 - 4.01) | 1.15 (1.08 - 1.23) | 3.74 (3.25 - 4.31) |
| 50s | 8.60 (8.10 - 9.13) | 1.03 (0.98 - 1.09) | 2.77 (2.63 - 2.93) | 3.70 (3.43 - 4.00) | 1.02 (0.94 - 1.11) | 4.11 (3.54 - 4.78) |
| 60s | 5.99 (5.57 - 6.44) | 0.77 (0.72 - 0.83) | 1.96 (1.82 - 2.10) | 2.80 (2.55 - 3.07) | 0.76 (0.67 - 0.85) | 3.57 (2.99 - 4.25) |
| 70s | 3.82 (3.47 - 4.21) | 0.74 (0.68 - 0.81) | 1.71 (1.55 - 1.87) | 2.18 (1.94 - 2.45) | 0.54 (0.45 - 0.64) | 2.67 (2.14 - 3.32) |
| ≥80 | 1.63 (1.39 - 1.91) | 0.84 (0.75 - 0.95) | 1.75 (1.55 - 1.97) | 2.74 (2.38 - 3.15) | 0.51 (0.40 - 0.65) | 1.88 (1.36 - 2.61) |
| Male | 0.88 (0.86 - 0.90) | 0.68 (0.66 - 0.69) | 1.07 (1.05 - 1.09) | 0.89 (0.86 - 0.92) | 1.27 (1.23 - 1.32) | 1.11 (1.05 - 1.17) |
| Obesity (BMI≥30) | 1.01 (0.96 - 1.07) | 0.83 (0.78 - 0.89) | 1.12 (1.06 - 1.18) | 1.53 (1.44 - 1.62) | 1.53 (1.41 - 1.65) | 1.33 (1.19 - 1.48) |
| Comorbidities |  |  |  |  |  |  |
| Malignancy | 0.96 (0.83 - 1.11) | 1.28 (1.10 - 1.48) | 1.17 (1.01 - 1.35) | 1.31 (1.11 - 1.53) | 0.72 (0.54 - 0.97) | 1.42 (1.07 - 1.88) |
| Immunocompromised | 1.27 (0.96 - 1.70) | 1.48 (1.11 - 1.97) | 1.23 (0.92 - 1.63) | 1.27 (0.93 - 1.73) | 1.50 (0.89 - 2.52) | 0.99 (0.55 - 1.79) |
| Chronic respiratory diseases | 1.43 (1.35 - 1.52) | 1.26 (1.19 - 1.34) | 1.50 (1.41 - 1.59) | 2.39 (2.25 - 2.54) | 1.41 (1.29 - 1.54) | 1.53 (1.36 - 1.72) |
| Chronic kidney diseases | 0.26 (0.11 - 0.61) | 1.05 (0.59 - 1.87) | 0.69 (0.39 - 1.23) | 1.15 (0.63 - 2.09) | 2.24 (1.07 - 4.69) | 0.44 (0.06 - 3.20) |
| Cardiovascular diseases | 1.21 (1.07 - 1.37) | 1.12 (0.99 - 1.28) | 1.36 (1.21 - 1.53) | 1.54 (1.35 - 1.75) | 1.15 (0.93 - 1.40) | 1.00 (0.76 - 1.32) |
| Cerebrovascular diseases | 1.03 (0.61 - 1.73) | 1.44 (0.88 - 2.36) | 1.08 (0.65 - 1.78) | 0.88 (0.48 - 1.62) | 2.42 (1.28 - 4.60) | 0.58 (0.14 - 2.36) |
| Hypertension | 0.99 (0.94 - 1.05) | 0.85 (0.80 - 0.91) | 0.94 (0.89 - 1.00) | 1.09 (1.02 - 1.17) | 1.17 (1.06 - 1.29) | 1.03 (0.91 - 1.17) |
| Diabetes | 1.01 (0.93 - 1.10) | 0.82 (0.74 - 0.90) | 1.05 (0.97 - 1.15) | 1.17 (1.07 - 1.29) | 1.14 (1.00 - 1.30) | 1.03 (0.86 - 1.22) |
| Vaccination ≥3 doses | 0.54 (0.52 - 0.55) | 0.39 (0.37 - 0.40) | 0.59 (0.58 - 0.61) | 0.90 (0.87 - 0.93) | 0.55 (0.52 - 0.57) | 0.70 (0.66 - 0.74) |
| Previous SARS-CoV-2 infection | 0.58 (0.54 - 0.62) | 0.46 (0.43 - 0.49) | 0.63 (0.59 - 0.67) | 1.04 (0.97 - 1.12) | 0.89 (0.81 - 0.98) | 0.83 (0.71 - 0.96) |

BMI, body mass index; CI, confidence interval; OR, odds ratio; SARS-CoV-2, severe acute respiratory syndrome coronavirus 2

**Table S4. Logistic regression analysis of factors for severe disease progression**

|  | Age < 65 | Age ≥ 65 |
| --- | --- | --- |
|  | Adjusted OR (95% CI) | Adjusted OR (95% CI) |
| Days from onset |  |  |
| 0 | (reference) | (reference) |
| 1 | 0.81 (0.31 - 2.15) | 0.74 (0.43 - 1.27) |
| 2 | 0.22 (0.05 - 0.88) | 0.79 (0.44 - 1.42) |
| 3 | 0.37 (0.09 - 1.54) | 0.66 (0.32 - 1.37) |
| 4 | 0.54 (0.10 - 2.79) | 0.23 (0.07 - 0.79) |
| 5 | 2.20 (0.57 - 8.52) | 1.23 (0.52 - 2.92) |
| BA.5 (compared with BA.2) | 0.93 (0.38 - 2.25) | 1.15 (0.61 - 2.17) |
| Age |  |  |
| <10 | (reference) | (reference) |
| 10s | 0.71 (0.13 - 4.03) | N/A |
| 20s | 0.29 (0.03 - 2.82) | N/A |
| 30s | 0.54 (0.09 - 3.27) | N/A |
| 40s | 1.82 (0.45 - 7.33) | N/A |
| 50s | 2.97 (0.72 - 12.32) | N/A |
| 60s | 7.92 (1.76 - 35.63) | (reference) |
| 70s | N/A | 2.14 (0.95 - 4.84) |
| ≥80 | N/A | 10.15 (4.70 - 21.92) |
| Male | 0.93 (0.44 - 1.96) | 1.61 (1.06 - 2.43) |
| Obesity (BMI≥30) | 3.55 (1.33 - 9.52) | 0.78 (0.21 - 2.92) |
| Comorbidities |  |  |
| Malignancy | 15.58 (4.88 - 49.80) | 2.61 (1.43 - 4.76) |
| Immunocompromised | 2.19 (0.35 - 13.65) | 0.38 (0.07 - 2.02) |
| Chronic respiratory diseases | 1.30 (0.35 - 4.75) | 2.92 (1.62 - 5.26) |
| Chronic kidney diseases | 28.34 (2.80 - 287.13) | 5.56 (1.53 - 20.16) |
| Cardiovascular diseases | 0.00 (0.00 - 0.00) | 1.01 (0.57 - 1.78) |
| Cerebrovascular diseases | 0.00 (0.00 - 0.00) | 4.23 (1.07 - 16.71) |
| Hypertension | 1.06 (0.33 - 3.38) | 0.57 (0.36 - 0.90) |
| Diabetes | 3.13 (0.96 - 10.20) | 1.39 (0.81 - 2.38) |
| Vaccination ≥3 doses | 0.36 (0.14 - 0.90) | 0.36 (0.24 - 0.56) |
| Previous SARS-CoV-2 infection | 0.77 (0.09 - 6.83) | 1.76 (0.37 - 8.44) |
| Symptoms |  |  |
| Fever | 0.79 (0.32 - 1.93) | 2.91 (1.89 - 4.51) |
| Cough | 1.86 (0.73 - 4.77) | 1.07 (0.68 - 1.69) |
| Sore throat | 0.41 (0.19 - 0.88) | 0.39 (0.24 - 0.63) |
| Nasal discharge | 0.91 (0.42 - 1.98) | 0.48 (0.28 - 0.82) |
| Phlegm | 1.16 (0.52 - 2.60) | 1.01 (0.63 - 1.63) |
| Headache | 0.66 (0.29 - 1.53) | 0.62 (0.31 - 1.22) |
| Joint or muscle pain≥ | 0.69 (0.28 - 1.71) | 0.55 (0.27 - 1.12) |
| Decreased food intake | 1.81 (0.79 - 4.14) | 2.41 (1.55 - 3.74) |
| Severe fatigue | 1.92 (0.83 - 4.42) | 1.93 (1.22 - 3.07) |
| Dyspnea | 2.78 (1.20 - 6.44) | 3.01 (1.84 - 4.91) |
| Diarrhea | 0.74 (0.20 - 2.76) | 0.93 (0.33 - 2.66) |
| Smell or taste disorder | 1.10 (0.24 - 5.02) | 6.5017826e-7 (0.00 - ∞) |

BMI, body mass index; CI, confidence interval; OR, odds ratio; SARS-CoV-2, severe acute respiratory syndrome coronavirus 2.

**Table S5. Logistic regression analysis of factors for symptoms in the group with three or more vaccination**

A. Fever, cough, sore throat, nasal discharge, phlegm, headache

|  | Fever | Cough | Sore throat | Nasal discharge | Phlegm | Headache |
| --- | --- | --- | --- | --- | --- | --- |
|  | Adjusted OR (95% CI) | | | | | |
| Months since last vaccination* | 1.10 (1.09 - 1.11) | 0.98 (0.97 - 0.99) | 0.98 (0.97 – 0.99) | 0.94 (0.93 – 0.95) | 1.01 (0.99 – 1.02) | 1.03 (1.02-1.04) |
| Days from onset |  | | | | | |
| 0 | (reference) | (reference) | (reference) | (reference) | (reference) | (reference) |
| 1 | 0.94 (0.89 - 0.99) | 1.41 (1.34 - 1.49) | 1.17 (1.10 - 1.24) | 1.28 (1.21 - 1.35) | 1.30 (1.23 - 1.38) | 1.09 (1.03 - 1.15) |
| 2 | 0.59 (0.56 - 0.63) | 2.15 (2.03 - 2.28) | 1.24 (1.17 - 1.31) | 1.79 (1.70 - 1.89) | 1.86 (1.75 - 1.97) | 1.00 (0.95 - 1.06) |
| 3 | 0.29 (0.27 - 0.31) | 2.16 (2.02 - 2.31) | 1.13 (1.06 - 1.21) | 2.03 (1.91 - 2.16) | 2.04 (1.91 - 2.17) | 0.75 (0.70 - 0.80) |
| 4 | 0.14 (0.12 - 0.16) | 1.94 (1.78 - 2.10) | 0.85 (0.79 - 0.92) | 1.79 (1.66 - 1.93) | 2.00 (1.85 - 2.16) | 0.56 (0.52 - 0.61) |
| 5 | 0.10 (0.08 - 0.12) | 1.62 (1.46 - 1.80) | 0.67 (0.60 - 0.74) | 1.61 (1.45 - 1.78) | 2.09 (1.89 - 2.31) | 0.49 (0.44 - 0.55) |
| BA.5 group  (compared with BA.2) | 2.28 (2.12 - 2.46) | 1.22 (1.16 - 1.30) | 1.10 (1.04 - 1.17) | 1.06 (1.00 - 1.11) | 1.09 (1.04 - 1.15) | 1.53 (1.44 - 1.61) |
| Age** |  |  |  |  |  |  |
| 10s | (reference) | (reference) | (reference) | (reference) | (reference) | (reference) |
| 20s | 0.90 (0.82 - 0.99) | 1.30 (1.17 - 1.43) | 0.92 (0.83 - 1.03) | 0.98 (0.90 - 1.08) | 1.51 (1.38 - 1.66) | 1.20 (1.09 - 1.32) |
| 30s | 0.80 (0.73 - 0.88) | 1.08 (0.98 - 1.19) | 0.79 (0.71 - 0.87) | 0.91 (0.84 - 1.00) | 1.28 (1.17 - 1.40) | 1.40 (1.28 - 1.53) |
| 40s | 0.75 (0.68 - 0.82) | 0.97 (0.88 - 1.06) | 0.73 (0.66 - 0.81) | 0.73 (0.67 - 0.80) | 1.05 (0.96 - 1.15) | 1.34 (1.22 - 1.46) |
| 50s | 0.65 (0.59 - 0.71) | 1.11 (1.00 - 1.22) | 0.71 (0.64 - 0.79) | 0.68 (0.62 - 0.74) | 0.86 (0.79 - 0.95) | 1.18 (1.07 - 1.29) |
| 60s | 0.46 (0.41 - 0.51) | 1.03 (0.93 - 1.14) | 0.57 (0.51 - 0.63) | 0.55 (0.50 - 0.61) | 0.74 (0.67 - 0.81) | 0.76 (0.69 - 0.84) |
| 70s | 0.35 (0.30 - 0.39) | 0.96 (0.86 - 1.07) | 0.38 (0.34 - 0.43) | 0.41 (0.37 - 0.45) | 0.64 (0.57 - 0.71) | 0.45 (0.40 - 0.51) |
| ≥80 | 0.35 (0.30 - 0.41) | 0.68 (0.60 - 0.77) | 0.16 (0.14 - 0.18) | 0.30 (0.26 - 0.34) | 0.57 (0.50 - 0.65) | 0.18 (0.15 - 0.22) |
| Male | 1.15 (1.11 - 1.20) | 0.84 (0.81 - 0.87) | 0.86 (0.83 - 0.89) | 0.87 (0.84 - 0.89) | 1.00 (0.97 - 1.04) | 0.62 (0.60 - 0.64) |
| BMI≥25 | 0.94 (0.86 - 1.02) | 1.36 (1.25 - 1.48) | 0.93 (0.86 - 1.01) | 1.58 (1.46 - 1.71) | 1.34 (1.24 - 1.44) | 0.98 (0.91 - 1.06) |
| Comorbidities |  |  |  |  |  |  |
| Malignancy | 1.34 (1.12 - 1.61) | 1.04 (0.88 - 1.22) | 0.92 (0.79 - 1.08) | 1.03 (0.88 - 1.19) | 1.05 (0.90 - 1.23) | 1.03 (0.87 - 1.22) |
| Immuno-compromised | 0.98 (0.68 - 1.41) | 0.96 (0.68 - 1.33) | 0.94 (0.68 - 1.29) | 1.01 (0.74 - 1.38) | 1.23 (0.90 - 1.69) | 1.08 (0.77 - 1.51) |
| Chronic respiratory diseases | 0.93 (0.84 - 1.03) | 1.56 (1.41 - 1.72) | 0.89 (0.81 - 0.97) | 1.39 (1.28 - 1.52) | 1.67 (1.54 - 1.82) | 1.33 (1.22 - 1.45) |
| Chronic kidney diseases | 1.06 (0.62 - 1.81) | 0.77 (0.49 - 1.21) | 0.79 (0.50 - 1.24) | 1.17 (0.74 - 1.84) | 0.80 (0.48 - 1.33) | 0.71 (0.40 - 1.28) |
| Cardiovascular diseases | 1.25 (1.08 - 1.46) | 1.21 (1.06 - 1.39) | 1.13 (0.99 - 1.29) | 1.18 (1.04 - 1.34) | 1.16 (1.02 - 1.32) | 0.95 (0.82 - 1.11) |
| Cerebrovascular diseases | 1.00 (0.53 - 1.88) | 0.81 (0.51 - 1.29) | 0.94 (0.59 - 1.48) | 0.87 (0.54 - 1.39) | 0.84 (0.51 - 1.39) | 0.66 (0.35 - 1.25) |
| Hypertension | 1.00 (0.93 - 1.08) | 1.08 (1.01 - 1.15) | 0.98 (0.92 - 1.04) | 1.06 (1.00 - 1.12) | 1.05 (0.99 - 1.11) | 0.90 (0.84 - 0.96) |
| Diabetes | 0.88 (0.79 - 0.97) | 1.09 (0.99 - 1.19) | 1.03 (0.95 - 1.12) | 1.14 (1.05 - 1.24) | 1.03 (0.95 - 1.13) | 0.94 (0.85 - 1.03) |
| Previous SARS-CoV-2 infection | 0.42 (0.35 - 0.50) | 0.72 (0.63 - 0.83) | 1.06 (0.91 - 1.23) | 0.99 (0.87 - 1.14) | 0.71 (0.61 - 0.81) | 0.72 (0.63 - 0.83) |

*Continuous predictor

**Patient group under 10 years of age was not applicable.

BMI, body mass index; CI, confidence interval; OR, odds ratio; SARS-CoV-2, severe acute respiratory syndrome coronavirus 2.

B. Joint or muscle pain, decreased food intake, severe fatigue, dyspnea, diarrhea, smell or taste disorder

|  | Joint or muscle pain | Decreased food intake | Severe fatigue | Dyspnea | Diarrhea | Smell or taste disorder |
| --- | --- | --- | --- | --- | --- | --- |
|  | Adjusted OR (95% CI) | | | | | |
| Months since last vaccination* | 1.07 (1.06 – 1.08) | 1.08 (1.07 – 1.10) | 1.07 (1.05 – 1.08) | 0.98 (0.96 – 0.99) | 1.01 (0.99 – 1.03) | 1.02 (0.99 – 1.05) |
| Days from onset |  | | | | | |
| 0 | (reference) | (reference) | (reference) | (reference) | (reference) | (reference) |
| 1 | 1.17 (1.10 - 1.24) | 1.13 (1.05 - 1.21) | 1.02 (0.96 - 1.09) | 1.12 (1.04 - 1.21) | 1.03 (0.91 - 1.16) | 0.94 (0.79 - 1.12) |
| 2 | 0.95 (0.89 - 1.01) | 1.02 (0.95 - 1.09) | 0.82 (0.77 - 0.87) | 1.35 (1.25 - 1.46) | 1.15 (1.02 - 1.30) | 1.33 (1.13 - 1.58) |
| 3 | 0.69 (0.64 - 0.74) | 0.80 (0.73 - 0.87) | 0.62 (0.57 - 0.66) | 1.28 (1.17 - 1.39) | 1.21 (1.06 - 1.39) | 2.13 (1.79 - 2.53) |
| 4 | 0.46 (0.42 - 0.51) | 0.61 (0.55 - 0.68) | 0.48 (0.43 - 0.53) | 1.17 (1.05 - 1.30) | 1.12 (0.94 - 1.33) | 2.90 (2.40 - 3.51) |
| 5 | 0.39 (0.34 - 0.45) | 0.53 (0.45 - 0.62) | 0.46 (0.40 - 0.52) | 1.11 (0.96 - 1.28) | 1.44 (1.17 - 1.77) | 4.55 (3.70 - 5.61) |
| BA.5 group  (compared with BA.2) | 1.56 (1.46 - 1.66) | 1.78 (1.64 - 1.93) | 1.54 (1.44 - 1.65) | 1.62 (1.50 - 1.75) | 1.21 (1.07 - 1.37) | 1.76 (1.49 - 2.08) |
| Age** |  |  |  |  |  |  |
| 10s | (reference) | (reference) | (reference) | (reference) | (reference) | (reference) |
| 20s | 1.65 (1.47 - 1.84) | 1.11 (0.99 - 1.25) | 1.51 (1.35 - 1.69) | 1.66 (1.47 - 1.88) | 1.68 (1.33 - 2.12) | 1.14 (0.89 - 1.47) |
| 30s | 1.99 (1.78 - 2.22) | 0.86 (0.77 - 0.97) | 1.62 (1.45 - 1.81) | 1.32 (1.17 - 1.50) | 1.84 (1.47 - 2.31) | 1.29 (1.01 - 1.65) |
| 40s | 1.95 (1.75 - 2.18) | 0.93 (0.83 - 1.04) | 1.35 (1.21 - 1.50) | 1.11 (0.98 - 1.26) | 1.76 (1.40 - 2.20) | 0.97 (0.76 - 1.25) |
| 50s | 1.76 (1.58 - 1.97) | 1.01 (0.90 - 1.13) | 1.16 (1.04 - 1.30) | 1.13 (1.00 - 1.29) | 1.40 (1.11 - 1.77) | 1.15 (0.89 - 1.47) |
| 60s | 1.23 (1.10 - 1.39) | 0.73 (0.65 - 0.83) | 0.83 (0.74 - 0.94) | 0.83 (0.73 - 0.95) | 1.11 (0.86 - 1.41) | 1.01 (0.78 - 1.31) |
| 70s | 0.83 (0.73 - 0.96) | 0.69 (0.60 - 0.80) | 0.74 (0.65 - 0.85) | 0.68 (0.58 - 0.79) | 0.74 (0.56 - 0.99) | 0.78 (0.58 - 1.06) |
| ≥80 | 0.38 (0.31 - 0.46) | 0.88 (0.75 - 1.04) | 0.80 (0.68 - 0.94) | 0.81 (0.68 - 0.97) | 0.73 (0.52 - 1.04) | 0.48 (0.32 - 0.73) |
| Male | 0.86 (0.82 - 0.89) | 0.48 (0.46 - 0.51) | 1.03 (0.99 - 1.07) | 0.90 (0.86 - 0.94) | 1.11 (1.03 - 1.20) | 0.95 (0.87 - 1.04) |
| BMI≥25 | 0.94 (0.87 - 1.03) | 0.76 (0.68 - 0.84) | 1.11 (1.02 - 1.20) | 1.57 (1.43 - 1.71) | 1.36 (1.18 - 1.57) | 1.42 (1.19 - 1.69) |
| Comorbidities |  |  |  |  |  |  |
| Malignancy | 0.94 (0.78 - 1.13) | 1.39 (1.15 - 1.68) | 1.22 (1.02 - 1.46) | 1.37 (1.13 - 1.67) | 0.74 (0.49 - 1.12) | 1.50 (1.05 - 2.16) |
| Immuno-compromised | 1.51 (1.07 - 2.15) | 1.51 (1.06 - 2.16) | 1.13 (0.79 - 1.61) | 1.28 (0.88 - 1.86) | 1.43 (0.71 - 2.88) | 0.95 (0.45 - 2.00) |
| Chronic respiratory diseases | 1.35 (1.23 - 1.47) | 1.19 (1.07 - 1.32) | 1.42 (1.29 - 1.55) | 2.42 (2.21 - 2.65) | 1.44 (1.23 - 1.69) | 1.47 (1.21 - 1.78) |
| Chronic kidney diseases | 0.33 (0.14 - 0.76) | 1.25 (0.67 - 2.34) | 0.87 (0.48 - 1.55) | 1.25 (0.69 - 2.28) | 2.63 (1.25 - 5.52) | 0.54 (0.07 - 3.91) |
| Cardiovascular diseases | 1.20 (1.03 - 1.40) | 1.21 (1.02 - 1.44) | 1.29 (1.11 - 1.49) | 1.54 (1.32 - 1.80) | 1.37 (1.05 - 1.78) | 1.08 (0.76 - 1.53) |
| Cerebrovascular diseases | 0.83 (0.43 - 1.62) | 1.29 (0.69 - 2.43) | 0.83 (0.44 - 1.59) | 0.69 (0.31 - 1.51) | 2.55 (1.16 - 5.60) | 0.39 (0.05 - 2.83) |
| Hypertension | 0.98 (0.91 - 1.05) | 0.88 (0.81 - 0.95) | 0.96 (0.90 - 1.04) | 1.08 (1.00 - 1.17) | 1.18 (1.03 - 1.34) | 1.01 (0.86 - 1.18) |
| Diabetes | 1.07 (0.97 - 1.18) | 0.90 (0.80 - 1.02) | 1.07 (0.97 - 1.19) | 1.17 (1.05 - 1.31) | 1.17 (0.98 - 1.40) | 0.93 (0.74 - 1.17) |
| Previous SARS-CoV-2 infection | 0.64 (0.54 - 0.75) | 0.60 (0.49 - 0.73) | 0.78 (0.66 - 0.92) | 0.89 (0.74 - 1.06) | 0.98 (0.74 - 1.30) | 0.78 (0.53 - 1.16) |

*Continuous variable

**Patient group under 10 years of age was not applicable.

BMI, body mass index; CI, confidence interval; OR, odds ratio. SARS-CoV-2, severe acute respiratory syndrome coronavirus 2.

**Table S6. COVID-19 symptoms frequency by days since symptom onset in total and subgroups**

A. Total individuals (N=157,861)

|  | Day 0 (onset)  (n=22,663) | Day 1  (n=52,383) | Day 2  (n=44,320) | Day 3  (n=22,927) | Day 4  (n=10,635) | Day 5  (n=4,933) |
| --- | --- | --- | --- | --- | --- | --- |
|  | Frequency (95% CI) | | | | | |
| Fever | 56.4 (55.7 - 57.0) | 52.8 (52.3 - 53.2) | 34.0 (33.5 - 34.4) | 18.3 (17.8 - 18.8) | 11.0 (10.4 - 11.6) | 7.9 (7.2 - 8.7) |
| Cough | 49.9 (49.2 - 50.5) | 58.6 (58.2 - 59.0) | 67.5 (67.1 - 68.0) | 70.9 (70.3 - 71.5) | 70.2 (69.3 - 71.1) | 69.0 (67.7 - 70.2) |
| Sore throat | 54.1 (53.4 - 54.7) | 60.0 (59.5 - 60.4) | 64.5 (64.1 - 65.0) | 64.7 (64.1 - 65.3) | 58.3 (57.3 - 59.2) | 51.6 (50.2 - 53.0) |
| Nasal discharge | 35.4 (34.8 - 36.0) | 40.0 (39.5 - 40.4) | 47.5 (47.0 - 47.9) | 51.9 (51.2 - 52.5) | 52.7 (51.7 - 53.6) | 50.3 (48.9 - 51.7) |
| Phlegm | 24.9 (24.4 - 25.5) | 31.5 (31.1 - 31.9) | 40.7 (40.2 - 41.1) | 43.8 (43.2 - 44.5) | 43.2 (42.3 - 44.2) | 42.9 (41.5 - 44.3) |
| Headache | 45.1 (44.4 - 45.7) | 46.9 (46.5 - 47.3) | 43.0 (42.6 - 43.5) | 35.8 (35.2 - 36.5) | 29.4 (28.5 - 30.2) | 25.3 (24.1 - 26.5) |
| Joint or muscle pain | 30.3 (29.7 - 30.9) | 33.8 (33.4 - 34.2) | 30.1 (29.7 - 30.5) | 23.8 (23.2 - 24.3) | 17.8 (17.1 - 18.5) | 14.2 (13.3 - 15.2) |
| Decreased food intake | 32.1 (31.5 - 32.7) | 32.9 (32.5 - 33.3) | 27.9 (27.5 - 28.3) | 21.6 (21.1 - 22.2) | 17.3 (16.6 - 18.0) | 14.4 (13.4 - 15.4) |
| Severe fatigue | 31.5 (30.9 - 32.1) | 31.3 (30.9 - 31.7) | 25.9 (25.5 - 26.3) | 20.5 (20.0 - 21.0) | 16.8 (16.1 - 17.6) | 14.9 (13.9 - 15.9) |
| Dyspnea | 12.5 (12.0 - 12.9) | 13.9 (13.6 - 14.2) | 16.6 (16.2 - 16.9) | 17.1 (16.6 - 17.6) | 16.5 (15.8 - 17.2) | 15.6 (14.6 - 16.6) |
| Diarrhea | 5.8 (5.5 - 6.1) | 7.3 (7.1 - 7.5) | 8.9 (8.6 - 9.1) | 8.8 (8.5 - 9.2) | 8.5 (8.0 - 9.1) | 8.4 (7.7 - 9.2) |
| Smell or taste disorder | 2.4 (2.2 - 2.6) | 2.6 (2.4 - 2.7) | 3.8 (3.6 - 4.0) | 5.2 (4.9 - 5.5) | 6.2 (5.8 - 6.7) | 9.5 (8.7 - 10.4) |

CI, confidence intervals.

B. Two or fewer vaccinations (N=99,389)

|  | Day 0 (onset)  (n=14,721) | Day 1  (n=33,651) | Day 2  (n=27,878) | Day 3  (n=13,837) | Day 4  (n=6,389) | Day 5  (n=2,913) |
| --- | --- | --- | --- | --- | --- | --- |
|  | Frequency -% (95% CI) | | | | | |
| Fever | 66.3 (65.5 - 67.1) | 61.7 (61.2 - 62.2) | 38.5 (38.0 - 39.1) | 20.8 (20.1 - 21.5) | 13.5 (12.7 - 14.4) | 10.0 (8.9 - 11.1) |
| Cough | 45.0 (44.2 - 45.8) | 53.8 (53.2 - 54.3) | 62.7 (62.2 - 63.3) | 67.7 (67.0 - 68.5) | 67.9 (66.8 - 69.0) | 68.4 (66.7 - 70.0) |
| Sore throat | 47.2 (46.4 - 48.0) | 53.9 (53.4 - 54.4) | 60.2 (59.6 - 60.8) | 61.7 (60.8 - 62.5) | 55.4 (54.2 - 56.6) | 48.8 (47.0 - 50.6) |
| Nasal discharge | 32.2 (31.5 - 33.0) | 35.6 (35.1 - 36.1) | 42.5 (41.9 - 43.1) | 47.6 (46.8 - 48.5) | 51.2 (50.0 - 52.4) | 49.6 (47.8 - 51.4) |
| Phlegm | 22.4 (21.7 - 23.1) | 29.2 (28.7 - 29.7) | 39.1 (38.6 - 39.7) | 43.2 (42.4 - 44.0) | 42.9 (41.6 - 44.1) | 41.9 (40.1 - 43.7) |
| Headache | 46.9 (46.1 - 47.7) | 48.3 (47.8 - 48.9) | 43.7 (43.1 - 44.3) | 36.7 (35.9 - 37.5) | 30.4 (29.3 - 31.5) | 26.1 (24.5 - 27.7) |
| Joint or muscle pain | 30.8 (30.1 - 31.6) | 34.3 (33.8 - 34.8) | 31.3 (30.8 - 31.9) | 25.2 (24.5 - 25.9) | 19.5 (18.5 - 20.5) | 15.3 (14.0 - 16.6) |
| Decreased food intake | 39.4 (38.6 - 40.2) | 39.9 (39.4 - 40.5) | 33.6 (33.0 - 34.1) | 26.1 (25.4 - 26.8) | 21.1 (20.1 - 22.1) | 17.5 (16.2 - 18.9) |
| Severe fatigue | 33.7 (32.9 - 34.4) | 33.1 (32.6 - 33.7) | 27.6 (27.1 - 28.1) | 22.1 (21.4 - 22.8) | 18.6 (17.7 - 19.6) | 16.1 (14.8 - 17.4) |
| Dyspnea | 11.6 (11.1 - 12.1) | 13.0 (12.6 - 13.3) | 15.8 (15.3 - 16.2) | 17.1 (16.5 - 17.8) | 17.3 (16.4 - 18.2) | 16.3 (15.0 - 17.7) |
| Diarrhea | 6.2 (5.9 - 6.6) | 8.5 (8.2 - 8.8) | 10.8 (10.5 - 11.2) | 10.9 (10.4 - 11.4) | 10.7 (10.0 - 11.5) | 9.8 (8.8 - 10.9) |
| Smell or taste disorder | 2.4 (2.2 - 2.6) | 2.7 (2.6 - 2.9) | 4.2 (3.9 - 4.4) | 5.4 (5.1 - 5.8) | 6.1 (5.5 - 6.7) | 9.6 (8.6 - 10.7) |

CI, confidence intervals.

C. Three or more vaccinations (N=58,472)

|  | Day 0 (onset)  (n=7,942) | Day 1  (n=18,732) | Day 2  (n=16,442) | Day 3  (n=9,090) | Day 4  (n=4,246) | Day 5  (n=2,020) |
| --- | --- | --- | --- | --- | --- | --- |
|  | Frequency -% (95% CI) | | | | | |
| Fever | 38.0 (36.9 - 39.0) | 36.7 (36.0 - 37.4) | 26.2 (25.5 - 26.9) | 14.4 (13.7 - 15.1) | 7.3 (6.5 - 8.1) | 5.0 (4.1 - 6.0) |
| Cough | 58.9 (57.8 - 60.0) | 67.2 (66.5 - 67.9) | 75.6 (75.0 - 76.3) | 75.7 (74.8 - 76.6) | 73.6 (72.3 - 74.9) | 69.8 (67.8 - 71.8) |
| Sore throat | 66.9 (65.8 - 67.9) | 70.8 (70.2 - 71.5) | 71.8 (71.1 - 72.5) | 69.4 (68.4 - 70.3) | 62.6 (61.1 - 64.0) | 55.7 (53.6 - 57.9) |
| Nasal discharge | 41.3 (40.2 - 42.4) | 47.8 (47.1 - 48.5) | 55.8 (55.1 - 56.6) | 58.4 (57.4 - 59.4) | 54.9 (53.4 - 56.4) | 51.4 (49.2 - 53.6) |
| Phlegm | 29.7 (28.7 - 30.7) | 35.5 (34.8 - 36.2) | 43.3 (42.5 - 44.0) | 44.9 (43.8 - 45.9) | 43.8 (42.3 - 45.3) | 44.3 (42.2 - 46.5) |
| Headache | 41.6 (40.6 - 42.7) | 44.3 (43.6 - 45.0) | 41.8 (41.1 - 42.6) | 34.5 (33.6 - 35.5) | 27.8 (26.5 - 29.2) | 24.2 (22.3 - 26.1) |
| Joint or muscle pain | 29.3 (28.3 - 30.3) | 32.9 (32.2 - 33.6) | 28.1 (27.4 - 28.8) | 21.6 (20.8 - 22.5) | 15.2 (14.2 - 16.3) | 12.7 (11.3 - 14.2) |
| Decreased food intake | 18.5 (17.6 - 19.3) | 20.2 (19.6 - 20.7) | 18.3 (17.7 - 18.9) | 14.8 (14.1 - 15.6) | 11.6 (10.6 - 12.6) | 9.9 (8.6 - 11.2) |
| Severe fatigue | 27.4 (26.5 - 28.4) | 27.9 (27.3 - 28.6) | 23.1 (22.5 - 23.8) | 18.0 (17.2 - 18.8) | 14.2 (13.2 - 15.3) | 13.2 (11.8 - 14.8) |
| Dyspnea | 14.1 (13.4 - 14.9) | 15.7 (15.1 - 16.2) | 18.0 (17.4 - 18.6) | 17.0 (16.2 - 17.8) | 15.4 (14.4 - 16.5) | 14.6 (13.1 - 16.2) |
| Diarrhea | 4.9 (4.5 - 5.4) | 5.1 (4.8 - 5.4) | 5.6 (5.2 - 5.9) | 5.7 (5.2 - 6.2) | 5.2 (4.5 - 5.9) | 6.4 (5.4 - 7.5) |
| Smell or taste disorder | 2.4 (2.1 - 2.8) | 2.3 (2.1 - 2.5) | 3.2 (2.9 - 3.5) | 4.9 (4.5 - 5.4) | 6.4 (5.7 - 7.2) | 9.5 (8.3 - 10.8) |

CI, confidence intervals.

D. No previous SARS-CoV-2 infection (N=152,094)

|  | Day 0 (onset)  (n=21,715) | Day 1  (n=50,464) | Day 2  (n=42,794) | Day 3  (n=22,117) | Day 4  (n=10,253) | Day 5  (n=4,751) |
| --- | --- | --- | --- | --- | --- | --- |
|  | Frequency -% (95% CI) | | | | | |
| Fever | 56.8 (56.2 - 57.5) | 53.3 (52.8 - 53.7) | 34.4 (34.0 - 34.9) | 18.4 (17.9 - 18.9) | 11.1 (10.5 - 11.8) | 8.0 (7.3 - 8.8) |
| Cough | 49.9 (49.2 - 50.6) | 58.6 (58.1 - 59.0) | 67.5 (67.1 - 68.0) | 70.9 (70.3 - 71.5) | 70.3 (69.4 - 71.2) | 68.6 (67.3 - 69.9) |
| Sore throat | 54.1 (53.4 - 54.7) | 60.1 (59.6 - 60.5) | 64.6 (64.1 - 65.0) | 64.8 (64.2 - 65.4) | 58.4 (57.4 - 59.3) | 51.6 (50.2 - 53.0) |
| Nasal discharge | 35.1 (34.4 - 35.7) | 39.6 (39.2 - 40.0) | 47.2 (46.7 - 47.7) | 51.7 (51.0 - 52.3) | 52.6 (51.7 - 53.6) | 50.1 (48.7 - 51.5) |
| Phlegm | 25.1 (24.5 - 25.7) | 31.5 (31.1 - 31.9) | 40.8 (40.4 - 41.3) | 44.0 (43.4 - 44.7) | 43.5 (42.5 - 44.5) | 43.0 (41.6 - 44.5) |
| Headache | 45.3 (44.6 - 45.9) | 47.2 (46.7 - 47.6) | 43.2 (42.7 - 43.7) | 35.9 (35.2 - 36.5) | 29.4 (28.5 - 30.2) | 25.2 (24.0 - 26.5) |
| Joint or muscle pain | 30.8 (30.2 - 31.4) | 34.1 (33.7 - 34.6) | 30.4 (30.0 - 30.9) | 23.9 (23.4 - 24.5) | 17.9 (17.2 - 18.7) | 14.2 (13.2 - 15.2) |
| Decreased food intake | 32.4 (31.8 - 33.0) | 33.2 (32.8 - 33.6) | 28.3 (27.9 - 28.7) | 21.9 (21.4 - 22.5) | 17.4 (16.7 - 18.2) | 14.6 (13.7 - 15.7) |
| Severe fatigue | 31.7 (31.1 - 32.4) | 31.6 (31.2 - 32.0) | 26.2 (25.8 - 26.6) | 20.6 (20.0 - 21.1) | 16.9 (16.2 - 17.6) | 15.0 (14.0 - 16.0) |
| Dyspnea | 12.5 (12.1 - 13.0) | 13.9 (13.6 - 14.2) | 16.6 (16.2 - 16.9) | 17.1 (16.6 - 17.6) | 16.4 (15.7 - 17.2) | 15.4 (14.4 - 16.4) |
| Diarrhea | 5.7 (5.4 - 6.0) | 7.3 (7.0 - 7.5) | 8.8 (8.6 - 9.1) | 8.8 (8.5 - 9.2) | 8.5 (8.0 - 9.1) | 8.4 (7.7 - 9.3) |
| Smell or taste disorder | 2.4 (2.2 - 2.6) | 2.6 (2.5 - 2.7) | 3.8 (3.6 - 4.0) | 5.2 (4.9 - 5.5) | 6.3 (5.8 - 6.8) | 9.7 (8.9 - 10.6) |

CI, confidence intervals.

E. Previous SARS-CoV-2 infection (N=5,767)

|  | Day 0 (onset)  (n=948) | Day 1  (n=1,919) | Day 2  (n=1,526) | Day 3  (n=810) | Day 4  (n=382) | Day 5  (n=182) |
| --- | --- | --- | --- | --- | --- | --- |
|  | Frequency -% (95% CI) | | | | | |
| Fever | 46.3 (43.2 - 49.5) | 39.7 (37.5 - 41.9) | 21.2 (19.2 - 23.3) | 15.7 (13.3 - 18.3) | 8.1 (5.8 - 11.3) | 4.9 (2.6 - 9.1) |
| Cough | 49.4 (46.2 - 52.5) | 58.4 (56.2 - 60.6) | 67.6 (65.2 - 69.9) | 71.1 (67.9 - 74.1) | 67.0 (62.2 - 71.5) | 78.6 (72.1 - 83.9) |
| Sore throat | 54.9 (51.7 - 58.0) | 57.0 (54.7 - 59.2) | 63.1 (60.7 - 65.5) | 62.0 (58.6 - 65.3) | 54.7 (49.7 - 59.6) | 52.7 (45.5 - 59.9) |
| Nasal discharge | 43.7 (40.5 - 46.8) | 48.8 (46.6 - 51.1) | 55.2 (52.7 - 57.7) | 57.5 (54.1 - 60.9) | 53.4 (48.4 - 58.3) | 55.5 (48.2 - 62.5) |
| Phlegm | 21.4 (18.9 - 24.1) | 29.6 (27.6 - 31.7) | 36.2 (33.8 - 38.6) | 39.1 (35.8 - 42.5) | 36.6 (32.0 - 41.6) | 38.5 (31.7 - 45.7) |
| Headache | 40.9 (37.8 - 44.1) | 39.7 (37.5 - 41.9) | 37.9 (35.5 - 40.3) | 35.2 (32.0 - 38.5) | 29.8 (25.5 - 34.6) | 26.4 (20.5 - 33.2) |
| Joint or muscle pain | 19.6 (17.2 - 22.3) | 24.4 (22.5 - 26.4) | 21.6 (19.6 - 23.7) | 20.0 (17.4 - 22.9) | 14.1 (11.0 - 18.0) | 14.3 (9.9 - 20.1) |
| Decreased food intake | 24.3 (21.6 - 27.1) | 23.7 (21.8 - 25.6) | 17.7 (15.9 - 19.7) | 13.6 (11.4 - 16.1) | 13.1 (10.1 - 16.8) | 7.7 (4.6 - 12.5) |
| Severe fatigue | 25.6 (23.0 - 28.5) | 23.3 (21.5 - 25.3) | 19.4 (17.5 - 21.5) | 18.8 (16.2 - 21.6) | 15.4 (12.2 - 19.4) | 13.2 (9.0 - 18.9) |
| Dyspnea | 11.2 (9.3 - 13.3) | 14.9 (13.3 - 16.5) | 17.4 (15.5 - 19.3) | 17.0 (14.6 - 19.8) | 19.1 (15.5 - 23.4) | 22.0 (16.6 - 28.5) |
| Diarrhea | 7.0 (5.5 - 8.8) | 7.9 (6.8 - 9.2) | 9.2 (7.9 - 10.8) | 8.4 (6.7 - 10.5) | 8.1 (5.8 - 11.3) | 7.1 (4.2 - 11.8) |
| Smell or taste disorder | 2.8 (2.0 - 4.1) | 2.1 (1.6 - 2.9) | 3.1 (2.4 - 4.1) | 5.9 (4.5 - 7.8) | 4.7 (3.0 - 7.3) | 5.5 (3.0 - 9.8) |

CI, confidence intervals.

**Table S7. Cox regression analysis for the presence of symptoms**

A. Fever, cough, sore throat, nasal discharge, phlegm, headache

|  | Fever | Cough | Sore throat | Nasal discharge | Phlegm | Headache |
| --- | --- | --- | --- | --- | --- | --- |
|  | Adjusted HR (95% CI) | | | | | |
| BA.5 group  (compared with BA.2) | 1.67 (1.64 - 1.71) | 1.08 (1.06 - 1.09) | 1.11 (1.09 - 1.13) | 0.96 (0.94 - 0.98) | 1.04 (1.02 - 1.06) | 1.36 (1.33 - 1.39) |
| Age |  |  |  |  |  |  |
| <10 | (reference) | (reference) | (reference) | (reference) | (reference) | (reference) |
| 10s | 0.82 (0.80 - 0.84) | 1.06 (1.03 - 1.09) | 2.51 (2.43 - 2.60) | 0.94 (0.91 - 0.96) | 1.82 (1.75 - 1.89) | 1.86 (1.79 - 1.92) |
| 20s | 0.68 (0.67 - 0.70) | 1.09 (1.07 - 1.12) | 2.52 (2.44 - 2.60) | 0.95 (0.92 - 0.97) | 2.27 (2.18 - 2.35) | 1.96 (1.90 - 2.03) |
| 30s | 0.66 (0.64 - 0.68) | 1.01 (0.98 - 1.03) | 2.36 (2.29 - 2.44) | 0.87 (0.84 - 0.89) | 2.03 (1.95 - 2.11) | 2.06 (1.99 - 2.13) |
| 40s | 0.59 (0.57 - 0.61) | 0.95 (0.93 - 0.97) | 2.25 (2.17 - 2.32) | 0.76 (0.74 - 0.79) | 1.79 (1.72 - 1.86) | 1.92 (1.86 - 1.99) |
| 50s | 0.51 (0.49 - 0.53) | 0.96 (0.93 - 0.99) | 2.16 (2.09 - 2.24) | 0.72 (0.69 - 0.74) | 1.59 (1.53 - 1.66) | 1.76 (1.70 - 1.83) |
| 60s | 0.35 (0.33 - 0.37) | 0.86 (0.83 - 0.89) | 1.80 (1.73 - 1.88) | 0.60 (0.57 - 0.62) | 1.29 (1.22 - 1.36) | 1.21 (1.15 - 1.27) |
| 70s | 0.24 (0.22 - 0.26) | 0.77 (0.74 - 0.81) | 1.41 (1.34 - 1.49) | 0.46 (0.43 - 0.49) | 1.08 (1.01 - 1.15) | 0.74 (0.69 - 0.79) |
| ≥80 | 0.30 (0.27 - 0.34) | 0.80 (0.76 - 0.85) | 1.06 (0.98 - 1.15) | 0.44 (0.40 - 0.47) | 1.15 (1.05 - 1.25) | 0.42 (0.37 - 0.48) |
| Male | 1.12 (1.11 - 1.14) | 0.98 (0.97 - 0.99) | 0.98 (0.97 - 0.99) | 0.98 (0.97 - 1.00) | 1.01 (0.99 - 1.03) | 0.88 (0.86 - 0.89) |
| Obesity (BMI≥30) | 1.01 (0.97 - 1.05) | 1.11 (1.07 - 1.14) | 0.97 (0.94 - 1.00) | 1.22 (1.18 - 1.26) | 1.19 (1.15 - 1.23) | 1.02 (0.98 - 1.06) |
| Comorbidities |  |  |  |  |  |  |
| Malignancy | 1.28 (1.15 - 1.43) | 1.05 (0.97 - 1.13) | 1.01 (0.94 - 1.10) | 1.01 (0.92 - 1.11) | 1.07 (0.97 - 1.19) | 1.00 (0.90 - 1.11) |
| Immuno-compromised | 1.01 (0.81 - 1.26) | 0.97 (0.83 - 1.13) | 0.99 (0.84 - 1.16) | 1.03 (0.86 - 1.25) | 1.13 (0.93 - 1.38) | 1.10 (0.90 - 1.34) |
| Chronic respiratory diseases | 1.05 (1.00 - 1.10) | 1.15 (1.12 - 1.19) | 1.01 (0.97 - 1.04) | 1.21 (1.16 - 1.25) | 1.32 (1.27 - 1.38) | 1.19 (1.15 - 1.24) |
| Chronic kidney diseases | 1.45 (0.92 - 2.27) | 1.42 (1.08 - 1.86) | 1.41 (1.05 - 1.90) | 1.69 (1.23 - 2.33) | 1.24 (0.82 - 1.88) | 1.02 (0.62 - 1.70) |
| Cardiovascular diseases | 1.17 (1.06 - 1.29) | 1.10 (1.04 - 1.17) | 1.09 (1.02 - 1.16) | 1.12 (1.04 - 1.21) | 1.13 (1.04 - 1.23) | 0.98 (0.89 - 1.08) |
| Cerebrovascular diseases | 1.12 (0.74 - 1.68) | 0.90 (0.71 - 1.15) | 0.92 (0.70 - 1.21) | 0.95 (0.70 - 1.30) | 0.86 (0.60 - 1.23) | 0.72 (0.45 - 1.14) |
| Hypertension | 1.04 (0.99 - 1.09) | 1.04 (1.01 - 1.07) | 1.01 (0.98 - 1.04) | 1.05 (1.01 - 1.09) | 1.05 (1.01 - 1.09) | 0.93 (0.90 - 0.98) |
| Diabetes | 0.88 (0.82 - 0.95) | 0.99 (0.95 - 1.04) | 0.96 (0.92 - 1.00) | 1.03 (0.98 - 1.08) | 0.99 (0.93 - 1.04) | 0.91 (0.85 - 0.96) |
| Vaccination ≥3 doses | 0.72 (0.71 - 0.73) | 1.19 (1.17 - 1.20) | 1.14 (1.12 - 1.16) | 1.43 (1.40 - 1.45) | 1.08 (1.06 - 1.10) | 0.85 (0.83 - 0.86) |
| Previous SARS-CoV-2 infection | 0.62 (0.59 - 0.65) | 1.02 (0.99 - 1.06) | 1.01 (0.98 - 1.05) | 1.21 (1.16 - 1.25) | 0.89 (0.85 - 0.94) | 0.84 (0.81 - 0.88) |

BMI, body mass index; CI, confidence interval; HR, hazard ratio; SARS-CoV-2, severe acute respiratory syndrome coronavirus 2.

The presence of symptoms on each day was considered as the appearance of symptoms and was set as the outcome of the Cox regression analysis.

B. Joint or muscle pain, decreased food intake, severe fatigue, dyspnea, diarrhea, smell or taste disorder

|  | Joint or muscle pain | Decreased food intake | Severe fatigue | Dyspnea | Diarrhea | Smell or taste disorder |
| --- | --- | --- | --- | --- | --- | --- |
|  | Adjusted HR (95% CI) | | | | | |
| BA.5 group  (compared with BA.2) | 1.47 (1.43 - 1.50) | 1.64 (1.60 - 1.68) | 1.55 (1.51 - 1.59) | 1.39 (1.34 - 1.44) | 1.27 (1.21 - 1.32) | 1.58 (1.47 - 1.70) |
| Age |  |  |  |  |  |  |
| <10 | (reference) | (reference) | (reference) | (reference) | (reference) | (reference) |
| 10s | 2.31 (2.19 - 2.44) | 0.94 (0.92 - 0.98) | 1.54 (1.48 - 1.61) | 2.32 (2.16 - 2.49) | 0.83 (0.78 - 0.89) | 2.57 (2.22 - 2.96) |
| 20s | 3.83 (3.64 - 4.02) | 0.81 (0.79 - 0.84) | 1.94 (1.87 - 2.02) | 3.62 (3.39 - 3.87) | 0.92 (0.87 - 0.98) | 3.64 (3.18 - 4.17) |
| 30s | 4.39 (4.18 - 4.61) | 0.74 (0.72 - 0.77) | 2.05 (1.97 - 2.14) | 2.96 (2.77 - 3.16) | 0.97 (0.91 - 1.03) | 3.55 (3.09 - 4.06) |
| 40s | 4.14 (3.94 - 4.35) | 0.73 (0.70 - 0.75) | 1.77 (1.70 - 1.85) | 2.54 (2.37 - 2.72) | 0.89 (0.84 - 0.95) | 2.91 (2.53 - 3.34) |
| 50s | 3.71 (3.52 - 3.92) | 0.74 (0.72 - 0.77) | 1.53 (1.46 - 1.61) | 2.46 (2.29 - 2.65) | 0.78 (0.72 - 0.85) | 3.11 (2.68 - 3.61) |
| 60s | 2.57 (2.41 - 2.74) | 0.53 (0.50 - 0.57) | 1.05 (0.98 - 1.11) | 1.75 (1.61 - 1.91) | 0.54 (0.48 - 0.60) | 2.48 (2.09 - 2.95) |
| 70s | 1.63 (1.49 - 1.77) | 0.46 (0.43 - 0.50) | 0.84 (0.77 - 0.92) | 1.30 (1.16 - 1.45) | 0.35 (0.30 - 0.42) | 1.75 (1.41 - 2.17) |
| ≥80 | 0.98 (0.84 - 1.13) | 0.63 (0.57 - 0.70) | 1.07 (0.96 - 1.19) | 1.83 (1.61 - 2.08) | 0.38 (0.30 - 0.48) | 1.37 (1.00 - 1.89) |
| Male | 0.97 (0.95 - 0.98) | 0.80 (0.79 - 0.82) | 1.10 (1.08 - 1.12) | 0.94 (0.92 - 0.97) | 1.29 (1.24 - 1.34) | 1.14 (1.08 - 1.20) |
| Obesity (BMI≥30) | 1.01 (0.97 - 1.06) | 0.88 (0.84 - 0.93) | 1.08 (1.03 - 1.13) | 1.39 (1.32 - 1.46) | 1.46 (1.36 - 1.57) | 1.31 (1.18 - 1.45) |
| Comorbidities |  |  |  |  |  |  |
| Malignancy | 0.98 (0.87 - 1.11) | 1.19 (1.05 - 1.35) | 1.13 (1.00 - 1.28) | 1.25 (1.08 - 1.44) | 0.74 (0.56 - 0.99) | 1.42 (1.08 - 1.86) |
| Immunocompromised | 1.18 (0.94 - 1.48) | 1.33 (1.06 - 1.67) | 1.18 (0.93 - 1.48) | 1.23 (0.94 - 1.61) | 1.48 (0.90 - 2.42) | 1.01 (0.57 - 1.80) |
| Chronic respiratory diseases | 1.27 (1.21 - 1.33) | 1.20 (1.15 - 1.26) | 1.34 (1.28 - 1.40) | 1.97 (1.87 - 2.07) | 1.39 (1.28 - 1.51) | 1.52 (1.35 - 1.70) |
| Chronic kidney diseases | 0.52 (0.23 - 1.15) | 1.54 (0.91 - 2.60) | 1.16 (0.69 - 1.97) | 1.66 (0.96 - 2.86) | 3.17 (1.58 - 6.37) | 0.66 (0.09 - 4.71) |
| Cardiovascular diseases | 1.17 (1.06 - 1.29) | 1.13 (1.01 - 1.26) | 1.30 (1.18 - 1.43) | 1.45 (1.30 - 1.62) | 1.16 (0.96 - 1.41) | 1.03 (0.79 - 1.35) |
| Cerebrovascular diseases | 1.04 (0.67 - 1.62) | 1.23 (0.81 - 1.88) | 1.03 (0.66 - 1.60) | 0.82 (0.47 - 1.45) | 2.29 (1.26 - 4.16) | 0.56 (0.14 - 2.25) |
| Hypertension | 1.00 (0.96 - 1.05) | 0.89 (0.85 - 0.95) | 0.97 (0.92 - 1.02) | 1.08 (1.02 - 1.15) | 1.16 (1.06 - 1.27) | 1.03 (0.91 - 1.17) |
| Diabetes | 0.97 (0.91 - 1.04) | 0.82 (0.75 - 0.89) | 1.00 (0.93 - 1.08) | 1.11 (1.02 - 1.20) | 1.10 (0.97 - 1.25) | 1.00 (0.84 - 1.19) |
| Vaccination ≥3 doses | 0.72 (0.70 - 0.73) | 0.53 (0.52 - 0.54) | 0.74 (0.72 - 0.76) | 0.96 (0.93 - 0.99) | 0.60 (0.57 - 0.62) | 0.74 (0.70 - 0.78) |
| Previous SARS-CoV-2 infection | 0.68 (0.65 - 0.72) | 0.56 (0.53 - 0.60) | 0.71 (0.67 - 0.75) | 1.01 (0.94 - 1.07) | 0.88 (0.80 - 0.96) | 0.82 (0.71 - 0.94) |

BMI, body mass index; CI, confidence interval; HR, hazard ratio; SARS-CoV-2, severe acute respiratory syndrome coronavirus 2.

The presence of symptoms on each day was considered as the appearance of symptoms and was set as the outcome of the Cox regression analysis.

**Table S8. Frequency of COVID-19 symptoms by age**

|  | Total  (N=157,552) | Age | | | | | | | | |
| --- | --- | --- | --- | --- | --- | --- | --- | --- | --- | --- |
|  |  | <10  (n=23,219) | 10s  (n=21,165) | 20s  (n=26,259) | 30s  (n=27,665) | 40s  (n=26,536) | 50s  (n=16,629) | 60s  (n=8,881) | 70s  (n=4,839) | 80 or over  (n=2,359) |
|  | Frequency -% (95% CI) | | | | | | | | | |
| Fever | 38.8 (38.5 - 39.0) | 54.3 (53.7 - 55.0) | 48.2 (47.5 - 48.8) | 41.2 (40.6 - 41.8) | 38.3 (37.7 - 38.9) | 34.1 (33.5 - 34.7) | 29.5 (28.8 - 30.2) | 20.6 (19.8 - 21.5) | 14.5 (13.6 - 15.6) | 16.5 (15.0 - 18.0) |
| Cough | 62.7 (62.5 - 63.0) | 46.7 (46.1 - 47.3) | 59.5 (58.8 - 60.1) | 69.6 (69.1 - 70.2) | 65.0 (64.5 - 65.6) | 63.8 (63.2 - 64.4) | 67.5 (66.8 - 68.2) | 69.5 (68.5 - 70.4) | 69.5 (68.1 - 70.7) | 61.6 (59.6 - 63.5) |
| Sore throat | 60.7 (60.5 - 60.9) | 22.0 (21.5 - 22.5) | 65.3 (64.7 - 66.0) | 73.4 (72.9 - 73.9) | 69.1 (68.5 - 69.6) | 67.7 (67.2 - 68.3) | 67.6 (66.9 - 68.3) | 63.5 (62.5 - 64.5) | 54.6 (53.2 - 56.0) | 35.1 (33.2 - 37.1) |
| Nasal discharge | 44.3 (44.1 - 44.6) | 36.8 (36.2 - 37.4) | 42.6 (41.9 - 43.2) | 50.6 (50.0 - 51.2) | 48.1 (47.5 - 48.7) | 44.6 (44.0 - 45.2) | 44.5 (43.8 - 45.3) | 43.8 (42.8 - 44.9) | 38.1 (36.8 - 39.5) | 30.8 (29.0 - 32.7) |
| Phlegm | 36.1 (35.8 - 36.3) | 15.7 (15.2 - 16.2) | 34.1 (33.4 - 34.7) | 47.6 (47.0 - 48.2) | 43.0 (42.4 - 43.6) | 39.3 (38.7 - 39.8) | 36.2 (35.5 - 37.0) | 33.4 (32.4 - 34.4) | 31.2 (29.9 - 32.5) | 28.1 (26.4 - 30.0) |
| Headache | 42.1 (41.8 - 42.3) | 21.2 (20.7 - 21.7) | 44.4 (43.7 - 45.0) | 50.2 (49.6 - 50.8) | 51.9 (51.3 - 52.4) | 48.8 (48.2 - 49.4) | 44.6 (43.8 - 45.4) | 32.0 (31.0 - 33.0) | 20.5 (19.4 - 21.7) | 10.1 (9.0 - 11.4) |
| Joint or muscle pain | 29.1 (28.9 - 29.3) | 8.2 (7.9 - 8.6) | 21.0 (20.5 - 21.6) | 36.3 (35.7 - 36.9) | 40.3 (39.7 - 40.9) | 38.0 (37.4 - 38.6) | 33.8 (33.1 - 34.6) | 24.1 (23.2 - 25.0) | 16.1 (15.1 - 17.1) | 8.4 (7.4 - 9.6) |
| Decreased food intake | 28.1 (27.9 - 28.3) | 34.7 (34.1 - 35.3) | 35.1 (34.5 - 35.8) | 30.4 (29.9 - 31.0) | 26.6 (26.0 - 27.1) | 25.3 (24.7 - 25.8) | 24.5 (23.9 - 25.2) | 16.8 (16.1 - 17.6) | 15.0 (14.0 - 16.1) | 18.4 (16.9 - 20.0) |
| Severe fatigue | 26.8 (26.5 - 27.0) | 15.0 (14.5 - 15.4) | 25.5 (24.9 - 26.0) | 33.5 (33.0 - 34.1) | 34.4 (33.8 - 34.9) | 29.8 (29.2 - 30.3) | 25.8 (25.1 - 26.5) | 18.3 (17.5 - 19.1) | 15.6 (14.6 - 16.7) | 17.4 (15.9 - 19.0) |
| Dyspnea | 15.1 (15.0 - 15.3) | 4.7 (4.4 - 5.0) | 12.7 (12.2 - 13.1) | 22.2 (21.7 - 22.7) | 18.3 (17.9 - 18.8) | 16.4 (16.0 - 16.8) | 16.7 (16.1 - 17.2) | 13.4 (12.7 - 14.1) | 11.3 (10.4 - 12.2) | 14.0 (12.7 - 15.5) |
| Diarrhea | 7.8 (7.7 - 8.0) | 8.3 (7.9 - 8.6) | 7.7 (7.3 - 8.0) | 8.8 (8.5 - 9.2) | 8.9 (8.6 - 9.3) | 8.2 (7.9 - 8.5) | 7.1 (6.7 - 7.5) | 5.0 (4.6 - 5.5) | 3.6 (3.1 - 4.2) | 3.3 (2.7 - 4.1) |
| Smell or taste disorder | 3.7 (3.7 - 3.8) | 1.1 (1.0 - 1.2) | 3.2 (3.0 - 3.5) | 5.0 (4.8 - 5.3) | 4.8 (4.5 - 5.0) | 4.0 (3.8 - 4.2) | 4.3 (4.0 - 4.7) | 3.8 (3.5 - 4.3) | 3.0 (2.6 - 3.6) | 2.0 (1.5 - 2.6) |

CI, confidence intervals.

**Table S9. Logistic regression analysis of factors for symptoms (set elderly as an explanatory variable)**

A. Fever, cough, sore throat, nasal discharge, phlegm, headache

|  | Fever | Cough | Sore throat | Nasal discharge | Phlegm | Headache |
| --- | --- | --- | --- | --- | --- | --- |
|  | Adjusted OR (95% CI) | | | | | |
| Days from onset |  | | | | | |
| 0 | (reference) | (reference) | (reference) | (reference) | (reference) | (reference) |
| 1 | 0.84 (0.81 - 0.87) | 1.43 (1.38 - 1.47) | 1.26 (1.22 - 1.30) | 1.21 (1.17 - 1.25) | 1.38 (1.33 - 1.43) | 1.06 (1.02 - 1.09) |
| 2 | 0.38 (0.37 - 0.40) | 2.09 (2.02 - 2.16) | 1.53 (1.48 - 1.59) | 1.64 (1.59 - 1.70) | 2.06 (1.99 - 2.14) | 0.91 (0.88 - 0.94) |
| 3 | 0.17 (0.16 - 0.17) | 2.42 (2.33 - 2.52) | 1.54 (1.48 - 1.60) | 1.96 (1.88 - 2.03) | 2.35 (2.26 - 2.45) | 0.68 (0.66 - 0.71) |
| 4 | 0.09 (0.09 - 0.10) | 2.33 (2.22 - 2.45) | 1.18 (1.13 - 1.24) | 2.04 (1.94 - 2.14) | 2.32 (2.21 - 2.44) | 0.52 (0.49 - 0.55) |
| 5 | 0.06 (0.06 - 0.07) | 2.19 (2.05 - 2.35) | 0.91 (0.86 - 0.97) | 1.87 (1.76 - 1.99) | 2.30 (2.16 - 2.46) | 0.43 (0.40 - 0.46) |
| BA.5 group  (compared with BA.2) | 2.13 (2.07 - 2.19) | 1.01 (0.99 - 1.04) | 1.09 (1.06 - 1.12) | 0.81 (0.79 - 0.83) | 0.96 (0.94 - 0.99) | 1.49 (1.45 - 1.53) |
| Elderly (age ≥65) | 0.44 (0.41 - 0.47) | 0.88 (0.84 - 0.92) | 0.50 (0.47 - 0.52) | 0.55 (0.53 - 0.58) | 0.64 (0.61 - 0.67) | 0.36 (0.34 - 0.38) |
| Male | 1.15 (1.12 - 1.18) | 0.85 (0.83 - 0.87) | 0.85 (0.83 - 0.86) | 0.91 (0.89 - 0.92) | 0.95 (0.93 - 0.97) | 0.72 (0.70 - 0.73) |
| Obesity (BMI≥30) | 0.94 (0.89 - 0.99) | 1.48 (1.40 - 1.56) | 1.16 (1.10 - 1.22) | 1.51 (1.43 - 1.59) | 1.57 (1.50 - 1.66) | 1.26 (1.20 - 1.33) |
| Comorbidities |  |  |  |  |  |  |
| Malignancy | 1.27 (1.10 - 1.47) | 1.11 (0.97 - 1.26) | 1.00 (0.88 - 1.14) | 0.91 (0.80 - 1.03) | 1.06 (0.93 - 1.21) | 1.01 (0.88 - 1.16) |
| Immuno-compromised | 1.01 (0.75 - 1.36) | 0.87 (0.66 - 1.14) | 0.98 (0.75 - 1.28) | 1.06 (0.81 - 1.37) | 1.24 (0.96 - 1.61) | 1.22 (0.93 - 1.59) |
| Chronic respiratory diseases | 1.01 (0.95 - 1.08) | 1.47 (1.38 - 1.56) | 0.97 (0.92 - 1.02) | 1.38 (1.31 - 1.46) | 1.58 (1.50 - 1.67) | 1.36 (1.29 - 1.44) |
| Chronic kidney diseases | 0.72 (0.42 - 1.22) | 0.77 (0.50 - 1.19) | 0.70 (0.45 - 1.07) | 1.08 (0.70 - 1.67) | 0.73 (0.45 - 1.20) | 0.49 (0.28 - 0.87) |
| Cardiovascular diseases | 1.08 (0.95 - 1.22) | 1.18 (1.05 - 1.32) | 1.04 (0.93 - 1.15) | 1.08 (0.97 - 1.20) | 1.12 (1.00 - 1.24) | 0.87 (0.77 - 0.97) |
| Cerebrovascular diseases | 1.03 (0.62 - 1.73) | 0.89 (0.59 - 1.34) | 0.81 (0.55 - 1.20) | 0.93 (0.62 - 1.40) | 0.83 (0.54 - 1.27) | 0.59 (0.35 - 0.99) |
| Hypertension | 0.85 (0.80 - 0.90) | 1.11 (1.06 - 1.17) | 0.98 (0.93 - 1.03) | 0.96 (0.91 - 1.01) | 0.99 (0.94 - 1.04) | 0.86 (0.82 - 0.91) |
| Diabetes | 0.79 (0.72 - 0.86) | 1.12 (1.04 - 1.21) | 1.00 (0.93 - 1.08) | 1.07 (1.00 - 1.15) | 0.99 (0.92 - 1.06) | 0.92 (0.85 - 0.99) |
| Vaccination ≥3 doses | 0.43 (0.41 - 0.46) | 1.12 (1.06 - 1.18) | 1.02 (0.96 - 1.07) | 1.53 (1.45 - 1.62) | 0.87 (0.82 - 0.92) | 0.73 (0.69 - 0.78) |
| Previous SARS-CoV-2 infection | 0.42 (0.41 - 0.43) | 1.65 (1.62 - 1.69) | 1.94 (1.90 - 1.99) | 1.75 (1.71 - 1.79) | 1.28 (1.25 - 1.31) | 0.90 (0.88 - 0.92) |

The difference from the analysis in Supplementary Table 5 is that the age factor is divided into 65 years and older or not.

BMI, body mass index; CI, confidence interval; OR, odds ratio; SARS-CoV-2, severe acute respiratory syndrome coronavirus 2.

B. Joint or muscle pain, decreased food intake, severe fatigue, dyspnea, diarrhea, smell or taste disorder

|  | Joint or muscle pain | Decreased food intake | Severe fatigue | Dyspnea | Diarrhea | Smell or taste disorder |
| --- | --- | --- | --- | --- | --- | --- |
|  | Adjusted OR (95% CI) | | | | | |
| Days from onset |  | | | | | |
| 0 | (reference) | (reference) | (reference) | (reference) | (reference) | (reference) |
| 1 | 1.16 (1.12 - 1.20) | 1.03 (0.99 - 1.06) | 0.98 (0.95 - 1.01) | 1.13 (1.08 - 1.19) | 1.29 (1.21 - 1.37) | 1.08 (0.98 - 1.19) |
| 2 | 0.99 (0.95 - 1.02) | 0.82 (0.79 - 0.85) | 0.77 (0.74 - 0.79) | 1.40 (1.34 - 1.47) | 1.63 (1.52 - 1.74) | 1.63 (1.48 - 1.80) |
| 3 | 0.72 (0.70 - 0.76) | 0.59 (0.57 - 0.62) | 0.57 (0.55 - 0.60) | 1.46 (1.39 - 1.54) | 1.66 (1.55 - 1.79) | 2.30 (2.08 - 2.56) |
| 4 | 0.51 (0.48 - 0.54) | 0.45 (0.42 - 0.48) | 0.46 (0.43 - 0.49) | 1.42 (1.33 - 1.52) | 1.62 (1.49 - 1.77) | 2.81 (2.50 - 3.16) |
| 5 | 0.40 (0.36 - 0.43) | 0.36 (0.33 - 0.40) | 0.40 (0.37 - 0.43) | 1.33 (1.22 - 1.45) | 1.61 (1.43 - 1.81) | 4.52 (3.97 - 5.13) |
| BA.5 group  (compared with BA.2) | 1.58 (1.53 - 1.62) | 1.80 (1.74 - 1.85) | 1.64 (1.59 - 1.69) | 1.38 (1.33 - 1.43) | 1.20 (1.15 - 1.26) | 1.55 (1.44 - 1.67) |
| Elderly (age ≥65) | 0.45 (0.42 - 0.47) | 0.77 (0.72 - 0.81) | 0.58 (0.55 - 0.61) | 0.63 (0.59 - 0.67) | 0.55 (0.49 - 0.61) | 0.69 (0.61 - 0.78) |
| Male | 0.86 (0.84 - 0.88) | 0.68 (0.66 - 0.69) | 1.05 (1.03 - 1.07) | 0.88 (0.85 - 0.90) | 1.27 (1.22 - 1.31) | 1.09 (1.04 - 1.15) |
| Obesity (BMI≥30) | 1.37 (1.30 - 1.44) | 0.82 (0.77 - 0.87) | 1.34 (1.27 - 1.42) | 1.75 (1.65 - 1.86) | 1.61 (1.48 - 1.74) | 1.52 (1.37 - 1.70) |
| Comorbidities |  |  |  |  |  |  |
| Malignancy | 1.07 (0.92 - 1.23) | 1.24 (1.07 - 1.44) | 1.18 (1.02 - 1.36) | 1.29 (1.10 - 1.52) | 0.70 (0.52 - 0.94) | 1.45 (1.09 - 1.92) |
| Immunocompromised | 1.31 (0.99 - 1.74) | 1.48 (1.11 - 1.97) | 1.27 (0.95 - 1.68) | 1.29 (0.94 - 1.75) | 1.53 (0.91 - 2.56) | 1.00 (0.55 - 1.82) |
| Chronic respiratory diseases | 1.46 (1.38 - 1.54) | 1.25 (1.18 - 1.33) | 1.50 (1.42 - 1.59) | 2.36 (2.23 - 2.51) | 1.41 (1.29 - 1.54) | 1.54 (1.37 - 1.74) |
| Chronic kidney diseases | 0.24 (0.11 - 0.56) | 1.04 (0.58 - 1.86) | 0.66 (0.37 - 1.17) | 1.12 (0.62 - 2.03) | 2.10 (1.00 - 4.37) | 0.42 (0.06 - 3.04) |
| Cardiovascular diseases | 1.16 (1.03 - 1.30) | 1.12 (0.98 - 1.27) | 1.34 (1.19 - 1.50) | 1.53 (1.35 - 1.74) | 1.10 (0.90 - 1.35) | 0.98 (0.74 - 1.29) |
| Cerebrovascular diseases | 1.00 (0.60 - 1.65) | 1.45 (0.88 - 2.37) | 1.06 (0.64 - 1.75) | 0.87 (0.47 - 1.60) | 2.30 (1.22 - 4.35) | 0.54 (0.13 - 2.22) |
| Hypertension | 1.06 (1.00 - 1.12) | 0.82 (0.77 - 0.88) | 0.89 (0.84 - 0.95) | 1.03 (0.97 - 1.10) | 1.10 (1.00 - 1.21) | 1.04 (0.92 - 1.17) |
| Diabetes | 1.07 (0.99 - 1.16) | 0.80 (0.72 - 0.88) | 1.03 (0.95 - 1.12) | 1.14 (1.04 - 1.24) | 1.11 (0.97 - 1.26) | 1.04 (0.87 - 1.23) |
| Vaccination ≥3 doses | 0.58 (0.54 - 0.62) | 0.46 (0.43 - 0.49) | 0.63 (0.59 - 0.67) | 1.03 (0.96 - 1.11) | 0.90 (0.81 - 0.99) | 0.81 (0.70 - 0.94) |
| Previous SARS-CoV-2 infection | 0.86 (0.84 - 0.89) | 0.37 (0.36 - 0.38) | 0.76 (0.74 - 0.77) | 1.10 (1.07 - 1.13) | 0.56 (0.53 - 0.58) | 0.85 (0.80 - 0.90) |

The difference from the analysis in Supplementary Table 5 is that the age factor is divided into 65 years and older or not.

BMI, body mass index; CI, confidence interval; OR, odds ratio; SARS-CoV-2, severe acute respiratory syndrome coronavirus 2.

**Table S10. Logistic regression analysis of factors (including symptom scores) for progression to severe disease**

|  | Age < 65 | Age ≥ 65 |
| --- | --- | --- |
|  | Adjusted OR (95% CI) | Adjusted OR (95% CI) |
| Days from onset |  |  |
| 0 | (reference) | (reference) |
| 1 | 0.82 (0.31 - 2.15) | 0.74 (0.43 - 1.27) |
| 2 | 0.23 (0.06 - 0.94) | 0.78 (0.43 - 1.39) |
| 3 | 0.46 (0.11 - 1.90) | 0.65 (0.32 - 1.34) |
| 4 | 0.71 (0.14 - 3.63) | 0.23 (0.07 - 0.77) |
| 5 | 2.98 (0.77 - 11.49) | 1.22 (0.51 - 2.88) |
| BA.5 (compared with BA.2) | 0.87 (0.36 - 2.11) | 1.13 (0.60 - 2.14) |
| Age |  |  |
| <10 | (reference) | (reference) |
| 10s | 0.71 (0.13 - 3.96) | N/A |
| 20s | 0.33 (0.03 - 3.09) | N/A |
| 30s | 0.59 (0.10 - 3.51) | N/A |
| 40s | 2.02 (0.52 - 7.92) | N/A |
| 50s | 3.52 (0.89 - 13.94) | N/A |
| 60s | 9.07 (2.06 - 39.86) | (reference) |
| 70s | N/A | 2.12 (0.94 - 4.78) |
| ≥80 | N/A | 10.15 (4.71 - 21.89) |
| Male | 0.91 (0.44 - 1.89) | 1.60 (1.06 - 2.42) |
| Obesity (BMI≥30) | 3.83 (1.42 - 10.33) | 0.77 (0.20 - 2.94) |
| Comorbidities |  |  |
| Malignancy | 14.68 (4.63 - 46.47) | 2.61 (1.43 - 4.76) |
| Immunocompromised | 2.33 (0.38 - 14.50) | 0.37 (0.07 - 2.00) |
| Chronic respiratory diseases | 1.52 (0.42 - 5.49) | 3.01 (1.68 - 5.39) |
| Chronic kidney diseases | 30.69 (3.06 - 307.82) | 5.56 (1.52 - 20.29) |
| Cardiovascular diseases | 0.00 (0.00 - 0.00) | 1.02 (0.58 - 1.79) |
| Cerebrovascular diseases | 0.00 (0.00 - 0.00) | 4.03 (1.00 - 16.27) |
| Hypertension | 1.10 (0.35 - 3.45) | 0.57 (0.36 - 0.91) |
| Diabetes | 3.15 (0.97 - 10.16) | 1.39 (0.81 - 2.38) |
| Vaccination ≥3 doses | 0.38 (0.15 - 0.95) | 0.38 (0.25 - 0.58) |
| Previous SARS-CoV-2 infection | 0.86 (0.10 - 7.45) | 1.81 (0.38 - 8.73) |
| Symptoms |  |  |
| Cough | 2.23 (0.89 - 5.61) | 1.09 (0.70 - 1.72) |
| Phlegm | 1.26 (0.56 - 2.83) | 1.04 (0.65 - 1.66) |
| Headache | 0.63 (0.28 - 1.44) | 0.62 (0.31 - 1.21) |
| Joint or muscle pain | 0.70 (0.29 - 1.70) | 0.55 (0.27 - 1.11) |
| Diarrhea | 0.81 (0.22 - 2.98) | 0.93 (0.33 - 2.66) |
| Smell or taste disorder | 1.17 (0.26 - 5.40) | 0.00 (0.00 - 0.00) |
| **Number of upper airway symptoms*** | | |
| 0 | (reference) | (reference) |
| 1 | 0.45 (0.19 - 1.06) | 0.41 (0.26 - 0.65) |
| 2 | 0.45 (0.16 - 1.24) | 0.20 (0.09 - 0.46) |
| **Number of systemic symptoms**** | | |
| 0 | (reference) | (reference) |
| 1 | 1.09 (0.34 - 3.45) | 2.98 (1.73 - 5.14) |
| 2 | 3.41 (1.21 - 9.58) | 7.46 (4.15 - 13.41) |
| 3 | 4.90 (1.52 - 15.80) | 14.38 (7.14 - 28.98) |
| 4 | 2.56 (0.27 - 24.62) | 40.72 (14.72 - 112.68) |

* “Sore throat” and “nasal discharge”

**“Fever”, “decreased food intake”, “severe fatigue”, and “dyspnea”

BMI, body mass index; CI, confidence interval; OR, odds ratio; SARS-CoV-2, severe acute respiratory syndrome coronavirus 2.

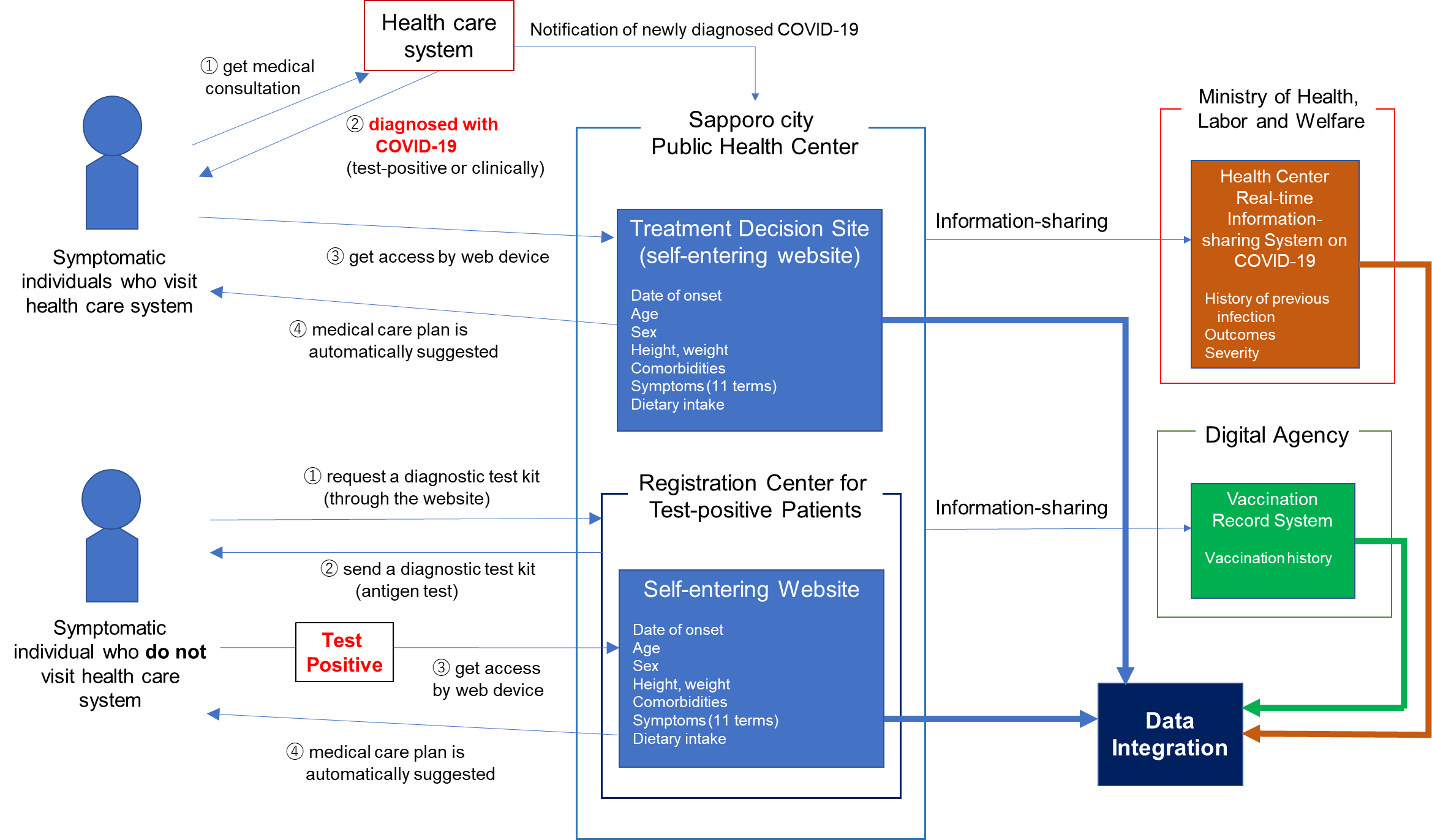

**Figure S1. Schema of information registration system for symptomatic COVID-19 patients in Sapporo.** Treatment Decision Site (TDS) is an online information registration system set up by Sapporo City Public Health Center. Registration Center for Test-positive Patients (RCPP) is a registration center operated by Sapporo City Public Health Center. The Ministry of Health, Labor and Welfare and Digital Agency are administrative agencies belonging to the Japanese government.

　　　
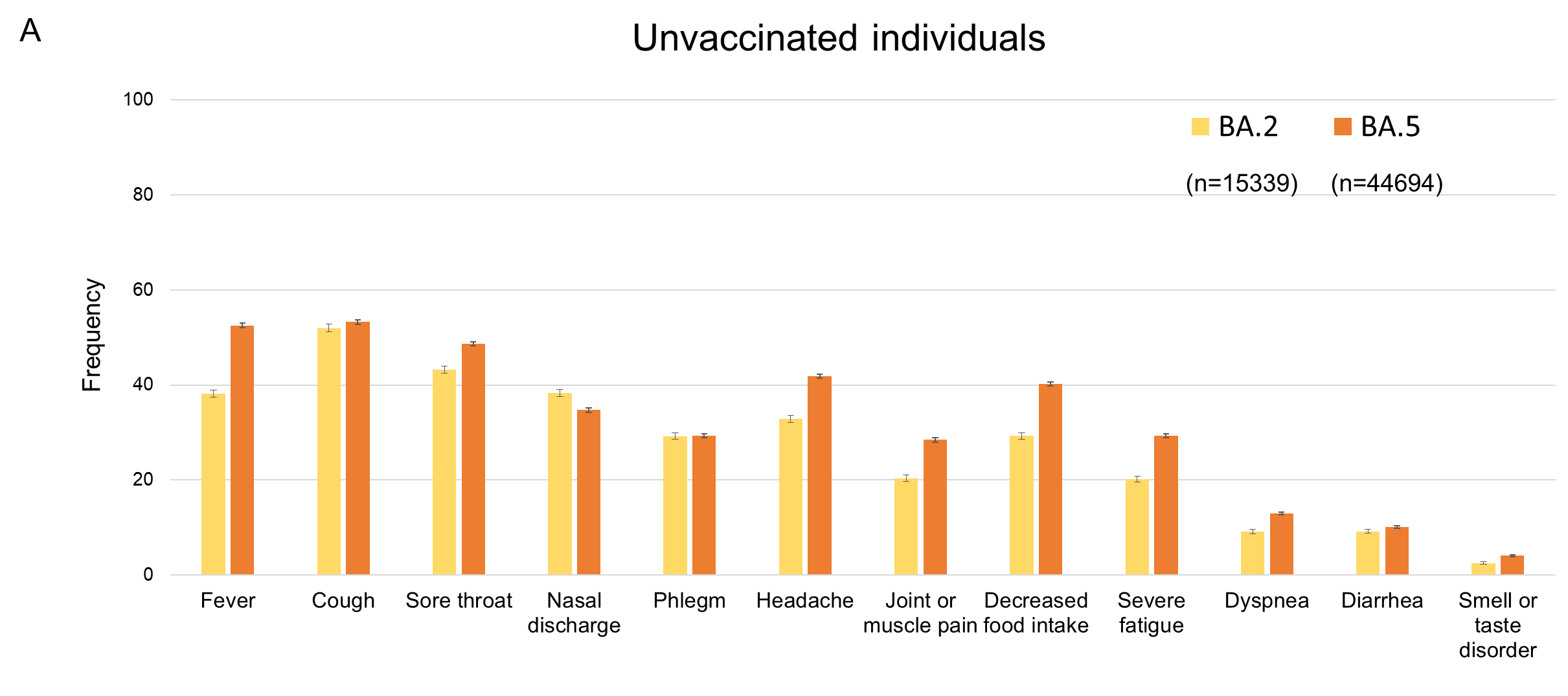

**Figure S2. COVID-19 symptom frequency in unvaccinated individuals.** The graph shows the frequency of 12 symptoms in unvaccinated individuals infected with omicron subvariants BA.2 or BA.5. Error bars indicate 95% confidence intervals.

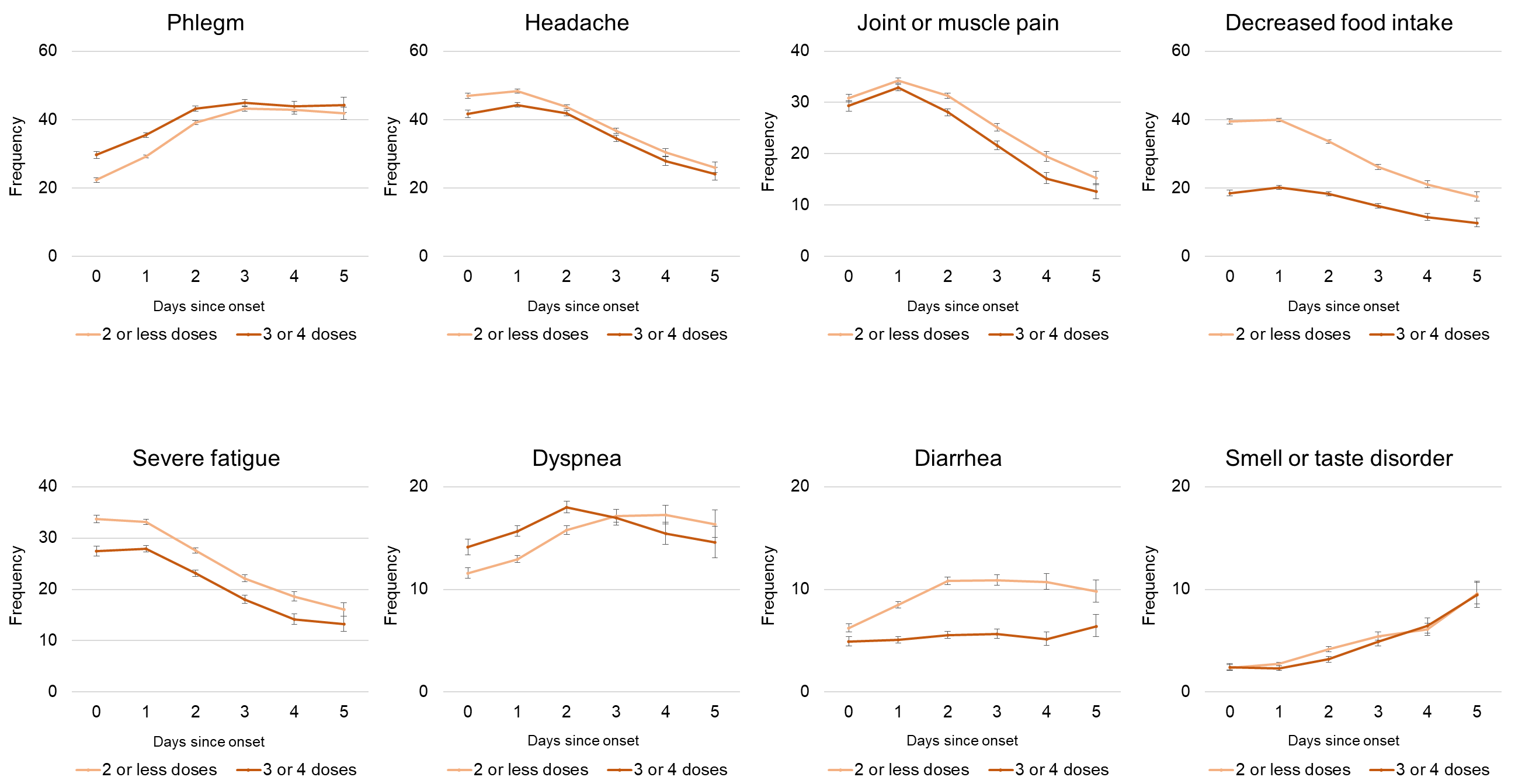

**Figure S3. Frequency of COVID-19 symptoms by days since symptom onset.** Individuals are divided into two subgroups according to vaccination status. Error bars indicate 95% confidence intervals. The frequency of fever, cough, sore throat and nasal discharge are shown in **Fig. 3**.

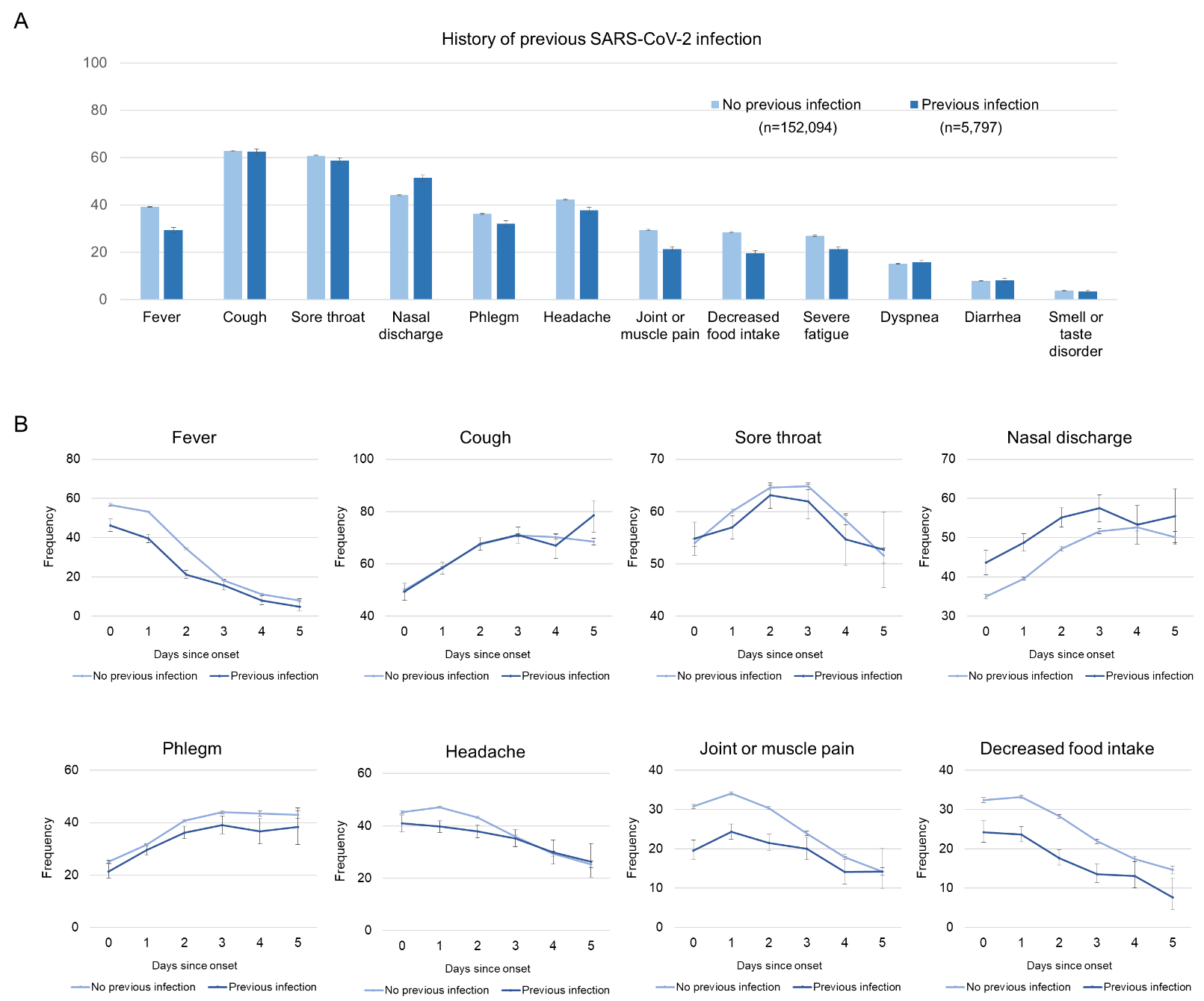
 (Figure continues on next page)

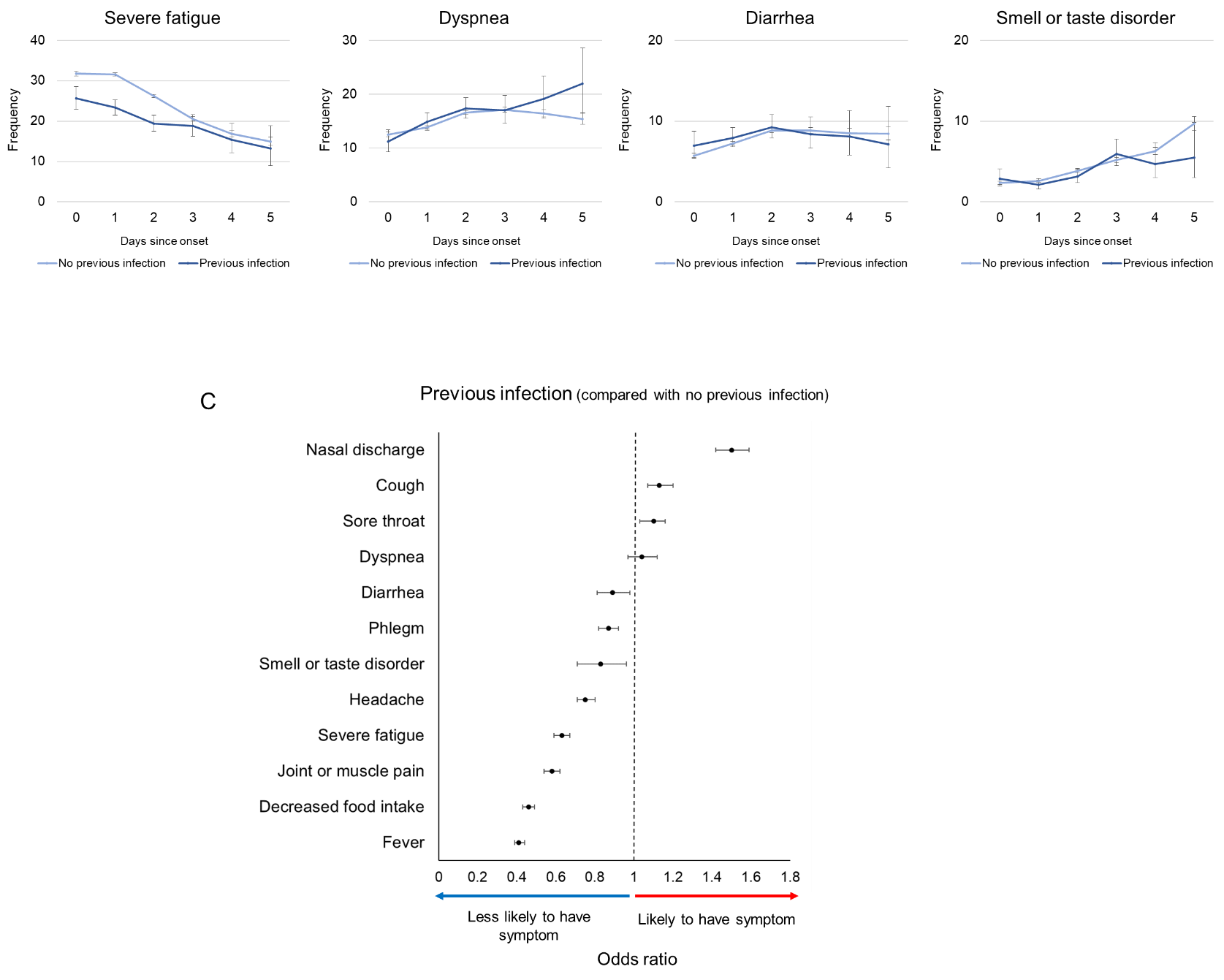

**Figure S4. Associations between history of previous infection and COVID-19 symptoms.** **A.** Frequency of COVID-19 symptoms in individuals with or without history of previous infection. **B.** Frequency of COVID-19 symptoms by days since symptom onset. **C.** Associations between previous COVID-19 infection and symptom likelihood. Multivariate analysis used each symptom as an outcome and mutant strain, age, body mass index, underlying disease, vaccination history, and history of spontaneous infection as explanatory variables. Among the explanatory variables, history of previous SARS-CoV-2 infection and the adjusted odds ratio (OR) for each symptom are arranged from highest to lowest. Points indicate odds ratios, and bars indicate 95% confidence intervals. Detailed results of the multivariate analysis are presented in **Table S3**.
